# Context-Specific Hypermetabolism In Humans With Mitochondrial Energy Transformation Defects

**DOI:** 10.64898/2026.05.30.26354528

**Authors:** Evan D Shaulson, Alexander J Sercel, Shannon Rausser, Samantha Leonard, Kathryn Whyte, Faris Zuraikat, Heather Seid, Qiuhan Huang, Alex Junker, Darshana Kapri, Mangesh Kurade, David Shire, Joanna Chen, Caroline Trumpff, Catherine Kelly, Natalia Bobba-Alves, Kris Englestad, Herman Pontzer, Wei Shen, Seth A Creasy, Edward L Melanson, Michael Rosenbaum, Marie-Pierre St-Onge, Dympna Gallagher, Michio Hirano, Martin Picard

## Abstract

Pathogenic mitochondrial DNA (mtDNA) defects provide an opportunity to test how impaired oxidative phosphorylation reshapes human energy expenditure. We studied adults with confirmed mtDNA defects (MitoD) and healthy controls using whole-room indirect calorimetry, doubly labeled water, quantitative magnetic resonance, actigraphy, autonomic monitoring, mood assessments, plasma inflammatory markers, and growth differentiation factor 15 (GDF15). Under behaviorally clamped conditions, fat-free mass-adjusted total energy expenditure (TEE) was consistently higher in MitoD, with higher non-resting expenditure per unit wrist acceleration by day, and blunted sleep-related metabolic suppression at night. In contrast, free-living TEE was similar between groups despite lower physical activity in MitoD, consistent with behavioral compensation within a constrained energy budget. Plasma GDF15 was ∼5-fold higher in MitoD and tracked with disease severity, systemic inflammation, fatigue, energetic burden, and lower habitual activity. These findings identify a persistent energetic burden in MitoD that is buffered in daily life and associated with elevated plasma GDF15.

---

Oxidative phosphorylation (OxPhos) is the primary mechanism by which human cells produce ATP and is central to human energy metabolism ^1^. As a result, genetic alterations that compromise OxPhos capacity offer a unique model to explore how mitochondrial energy transformation influence whole-body energy expenditure. Consistent with classic ^2^ and recent ^3,4^ reports of hypermetabolism in humans with OxPhos defects, we recently found that OxPhos-deficient human fibroblasts synthesize ATP at almost twice the rate (+91-101%) of control cells, despite slower cell division ^4^. In mice, genetic mitochondrial defects can strongly influence total energy expenditure (TEE) ^5^; some loss-of-function mutations significantly increase ^6–10^ while others significantly decrease mass-adjusted TEE ^11–14^. Accordingly, pathogenic mitochondrial DNA (mtDNA) defects provide a natural opportunity for testing how impaired cellular energy transformation reshapes whole-body energy expenditure in humans.

Among adults, pathogenic mtDNA defects include the m.3243A>G point mutation and single large-scale deletions, which underlie the most common mitochondrial disease (MitoD) syndromes such as mitochondrial encephalomyopathy, lactic acidosis, stroke-like episodes (MELAS) ^15^, chronic progressive external ophthalmoplegia (CPEO), and Kearns-Sayres Syndrome (KSS) ^16^. These disorders are marked by fatigue ^17^, physical inactivity ^18^, autonomic abnormalities ^19^, circulating immune-related changes ^20^, and exaggerated exercise-induced cardiorespiratory responses associated with impaired oxygen extraction and exercise intolerance ^21–23^. Several of these features could raise the energetic cost of daily life ^5^. Others, especially lower habitual physical activity due to severe fatigue, could lower it. No study has rigorously examined this question in MitoD using state-of-the-art measures of TEE ^24^.

Resolving the net effect of these opposing pressures on energy expenditure in MitoD requires measurements of metabolic rate in different contexts. First, measuring resting energy expenditure (REE) for 1-h under early morning fasted conditions minimizes the contribution of movement, posture, stress, feeding, and sleep on metabolic rate ^25^. Second, measuring TEE by whole-room indirect calorimetry (WRIC) captures the energy costs that the REE measurement assumes to be fixed and/or minimal (activity, feeding, etc.) while standardizing behavior ^26^. Third, doubly-labeled water (DLW)-based TEE measurement removes the constraints imposed by standardizing behavior, especially physical activity, so that energy expenditure (EE) can be estimated under habitual conditions that integrate subjective experiences (e.g., fatigue) ^27^. Combining these three methods within the same individuals allows disease-associated energy costs to be characterized in terms of the conditions under which they appear. Conceptually, one framework that may help interpret differences between these contexts is the constrained model of energy expenditure, which remains controversial, positing that TEE is maintained within a narrow range by competitive tradeoffs between physical activity and resting expenditures ^28–32^.

The cellular and physiological mechanisms by which subcellular OxPhos defects influence behavior, subjective experience, and systemic energy metabolism remain mostly unclear. However, converging lines of evidence point to growth differentiation factor-15 (GDF15), a robust biomarker of MitoD severity ^20,33–35^, as a candidate body-to-brain signal linking mitochondrial stress to organismal responses ^36^. Consistent with this notion, we recently found in human fibroblasts that selective OxPhos deficits rapidly cause reductive stress and chronically activate the integrated stress response (ISR) ^4^, a pathway mechanistically linked to GDF15 secretion ^37^. Signaling through its brainstem restricted receptor GDNF family receptor alpha-like (GFRAL) ^36,38–40^, GDF15 induces aversive visceral malaise states ^41^, suppresses appetite ^42^, reduces physical activity ^43,44^, promotes lean mass loss ^45^, and increases sympathetic nervous system (SNS) outflow ^46,47^. In humans with cancer cachexia, neutralizing GDF15 significantly improves appetite, body weight, and physical activity ^48^. These findings position GDF15 as a candidate body-to-brain signal ^36^ through which mitochondrial defects may cause fatigue, reduce physical activity, and drive the autonomic dysregulation that characterize MitoD.

Integrating these organelle-to-organism concepts, we applied WRIC and DLW to measure TEE and its components in adults with genetically confirmed pathogenic mtDNA defects and healthy controls. The 11-day protocol (**Figure 1**) included rigorous measures of body composition, continuous measurements of physical activity, sleep, and autonomic function, together with hourly measures of mood, plasma inflammatory markers and GDF15. Our findings reveal that under standardized WRIC conditions, TEE was significantly elevated in MitoD, driven by higher energy cost per unit movement during waking hours, and blunted metabolic suppression during sleep. In contrast, free-living TEE measured by DLW was similar between groups, accompanied by persistently lower physical activity and heart rate variability (HRV) in MitoD. Plasma GDF15, as well as the inflammatory marker c-reactive protein (CRP), were markedly elevated in MitoD compared to controls; they also correlated with fatigue, movement inefficiency, and reduced physical activity. These multi-level findings provide an integrated energetic account of mitochondrial disease pathophysiology and highlight a broader principle around the methodological importance of measuring energy expenditure across complementary contexts.

**Figure 1.**
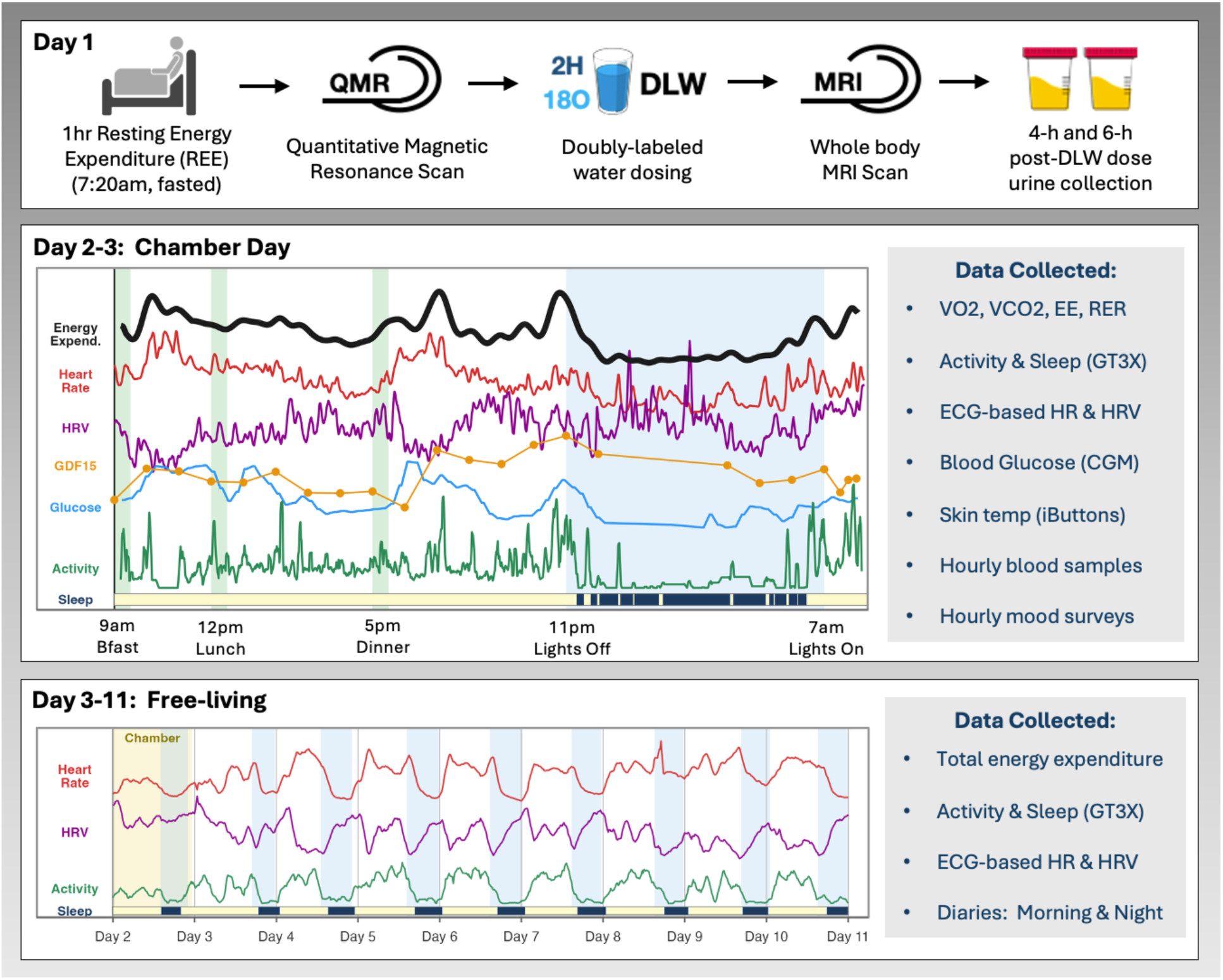
Mitochondrial Daily Energy Expenditure (MDEE) Study protocol overview. The schematic summarizes the 11-day MDEE study design. On Day 1, participants underwent fasting resting energy expenditure measurement by whole-room indirect calorimetry (WRIC), quantitative magnetic resonance body composition, doubly labeled water dosing, whole-body MRI, and 4-h and 6-h post-dose urine collections. On Days 2–3, participants completed a 23-h stay in the WRIC with standardized meals, scheduled lights-on and lights-off periods, wrist actigraphy, ECG-based heart rate and heart-rate variability monitoring, continuous glucose monitoring, skin temperature monitoring, hourly blood sampling, and hourly mood surveys. During Days 3–11, participants were studied under free-living conditions with doubly labeled water-based total energy expenditure, wrist actigraphy, ECG-based heart rate and heart-rate variability monitoring, and morning and evening diaries. Representative traces illustrate the time-resolved data streams collected during the chamber and free-living phases. *Abbreviations*: DLW, doubly labeled water; ECG, electrocardiography; MRI, magnetic resonance imaging; QMR, quantitative magnetic resonance; REE, resting energy expenditure; WRIC, whole-room indirect calorimetry.

## RESULTS

### Subject demographics, anthropometrics and body composition

To study the effects of mtDNA defects on energy expenditure and allocation, we compared adults with genetically confirmed pathogenic mtDNA variants (MitoD; n=10) with healthy controls (n=10) of similar age, sex, height, weight, and body mass index (**Figure 2A**).

**Figure 2:**
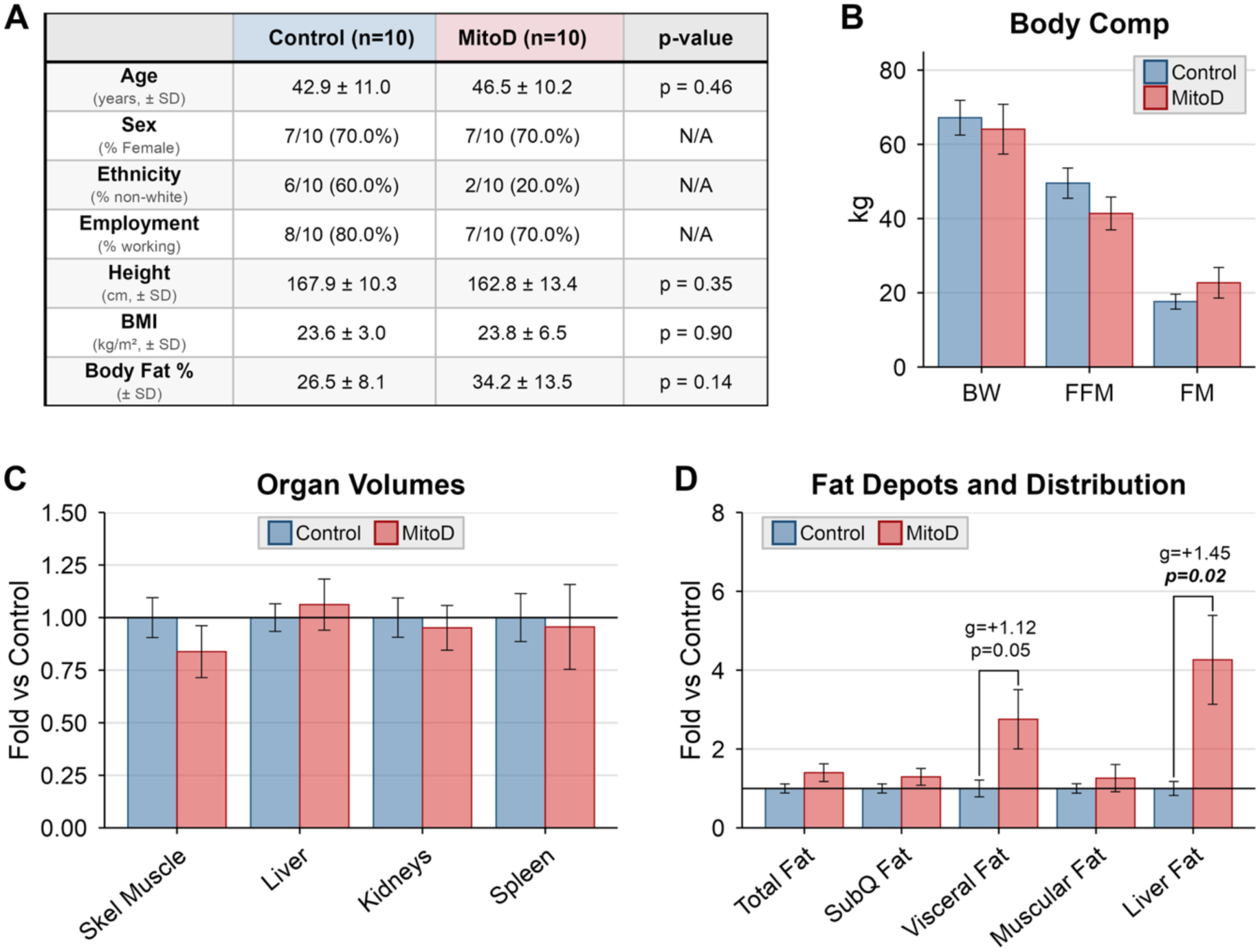
Cohort characteristics, body composition, organ volumes, and fat distribution. (A) Cohort characteristics, including age, sex, ethnicity, employment status, height, body mass index (BMI), and body fat percentage. (B) Body weight (BW) and fat mass (FM) measured by quantitative magnetic resonance., Fat-free mass (FFM) derived from the difference between body weight and fat mass. (C) MRI-derived skeletal muscle, liver, kidneys, and spleen volumes expressed relative to the control mean. (D) MRI-derived total adipose tissue, subcutaneous adipose tissue, visceral adipose tissue, intramuscular adipose tissue, and liver fat fraction expressed relative to the control mean. In panel A, continuous variables are mean ± SD and categorical variables are n/10 (%). In panels B–D, bars show mean ± SEM. Between-group p values are from Welch’s two-sided t-tests and are shown where indicated on the panels. *Abbreviations*: BMI, body mass index; BW, body weight; FFM, fat-free mass; FM, fat mass; MitoD, mitochondrial DNA-defect group; MRI, magnetic resonance imaging.

Body composition measured by quantitative magnetic resonance (QMR) showed no significant between-group differences in fat-free mass (FFM; MitoD 16% lower, p=0.19) or fat mass (FM; MitoD 29% higher, p=0.29) (**Figure 2B**). Magnetic resonance imaging (MRI) likewise showed similar volumes of skeletal muscle and major metabolically active organs, including the liver, kidneys, and spleen (**Figure 2C; Extended Data figure 1A**). Total adipose tissue, subcutaneous adipose tissue, and intramuscular adipose tissue volumes were also similar between groups, whereas visceral adipose tissue tended to be higher in MitoD (+176% higher, p=0.054), and liver fat fraction was significantly elevated (+326% higher, p=0.023) (**Figure 2D**; **Extended Data figure 1B**). Together, these data indicate that the groups were broadly similar in terms of overall body size, with differences emerging mainly in visceral adiposity and liver fat.

### Elevated TEE under controlled WRIC conditions despite similar REE

We used WRIC to quantify TEE under standardized conditions across the 23-h chamber stay. After adjusting for FFM using allometric residuals ^49^, TEE was significantly higher in MitoD than controls (residual TEE in MitoD: +195.5 kcal/day; p=0.016, **Figure 3A**). The group difference in TEE was of similar magnitude and significance during both the lights-on period (p=0.034) and the lights-off period (p=0.019, **Figure 3C**), indicating consistent hypermetabolism across day and night.

**Figure 3:**
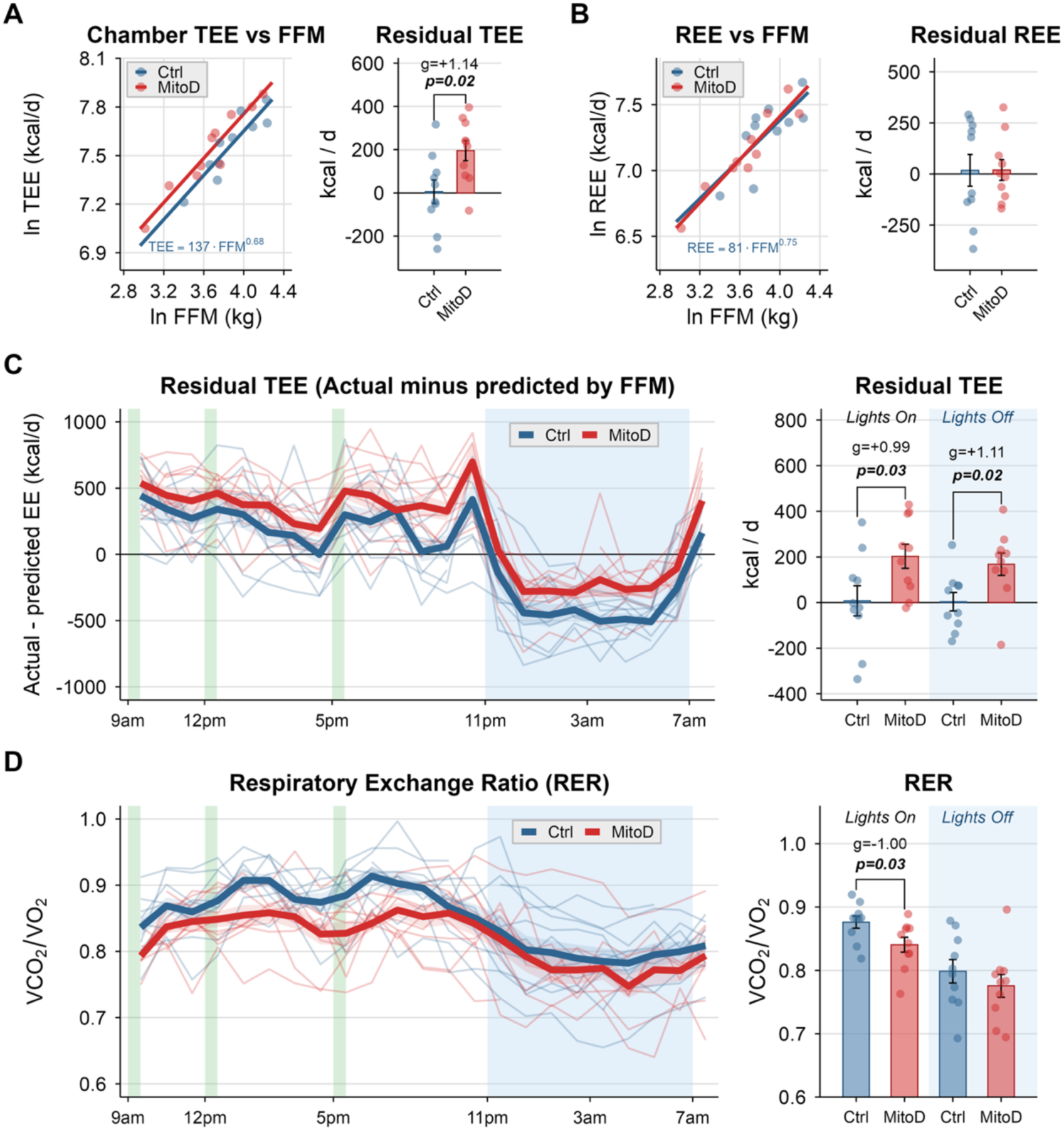
Chamber total energy expenditure and substrate utilization. (A) Natural-log-transformed chamber total energy expenditure (TEE) plotted against natural-log-transformed fat-free mass (FFM), with adjacent FFM-adjusted residual TEE values derived from the control allometric model. (B) Natural-log-transformed resting energy expenditure (REE) plotted against natural-log-transformed FFM, with adjacent FFM-adjusted residual REE values derived from the control allometric model. (C) Residual chamber TEE (actual minus predicted by FFM) across the 23-h chamber day, with thin lines showing individuals and thick lines showing group means; period means for the lights-on and lights-off intervals are shown at right. (D) Respiratory exchange ratio (RER) across the chamber day, with thin lines showing individuals and thick lines showing group means; period means for the lights-on and lights-off intervals are shown at right. Green vertical bands indicate standardized meals and blue shading indicates the lights-off interval. In bar panels, data are mean ± SEM with individual values. Between-group p values are from Welch’s two-sided t-tests. *Abbreviations*: FFM, fat-free mass; REE, resting energy expenditure; RER, respiratory exchange ratio; TEE, total energy expenditure.

To ensure the robustness of the elevated WRIC TEE finding given our small sample size and trend towards differences in body composition, we compared allometric TEE residuals using two additional normalization approaches: (1) an ANCOVA model adjusting for both FFM and FM, fit to our control subjects [ln(TEE) ∼ ln(FFM) + ln(FM)] ^49^, and (2) a published allometric equation derived from thousands of adults aged 20-60 from the DLW database of the International Atomic Energy Agency (see Extended Data figure 2 of ^50^). The magnitude and significance of MitoD’s TEE elevation were consistent across all three approaches (p=0.016-0.020; **Extended Data figure 2A**).

In contrast, REE, measured separately in the same room calorimeter for 1-h under tightly controlled early morning fasted conditions, did not differ between groups after the same FFM adjustment (residual REE in MitoD: +19.3 kcal/day; p=0.989, **Figure 3B**). A similar pattern was observed in the larger parent MiSBIE cohort, in which REE, measured under less stringent conditions (seated upright posture with VO_2_-only REEvue device) also did not differ between groups in either the fasted morning condition (residual REE in MitoD: +64.2 kcal/day; p=0.58) or fed afternoon condition (residual REE in MitoD: +118.8 kcal/day; p=0.21, **Extended Data figure 2B**). This finding localizes the group difference in TEE to energy costs captured over the standardized 23-h WRIC day, but not during short-term (1-h) tightly controlled resting measurement.

Mean respiratory exchange ratio (RER) did not differ between groups during the lights-off period. However, during the lights-on period, mean RER was significantly lower in MitoD than controls (−4.1%; p=0.031) and the typical postprandial rise in RER ^51^ appeared to be attenuated relative to controls (**Figure 3D**). This distinct daytime RER pattern may reflect a shift toward fat oxidation under fed conditions but could also be an artifact of negative energy balance ^52^, as the MitoD group’s energy intake during the WRIC day (from compulsory meals with energy content based on their predicted TEE) did not fully match their actual TEE (95.1% versus 107.5% in controls, **Extended Data figure 2D**).

### Increased expenditure per unit acceleration

To determine whether the elevated WRIC TEE observed in MitoD could be explained by greater spontaneous physical activity, we analyzed wrist accelerometry across the 23-h WRIC period. Mean acceleration, quantified as Euclidean Norm Minus One (ENMO) was lower in MitoD (−26%; p=0.019), as was peak 30-min average ENMO (−35%; p=0.001). MitoD participants also spent less time in moderate-to-vigorous physical activity (Control 9.5 ± 7.1 vs MitoD 2.7 ± 3.2 min/d; p=0.016) and light activity (Control 132.0 ± 52.7 min/d vs MitoD 62.0 ± 36.1 min/d p=0.002) with more time spent inactive (Control 803.5 ± 71.4 vs MitoD 909.1 ± 54.8 min/d; p=0.002, **Figure 4A**). Thus, the elevated WRIC TEE in MitoD relative to controls cannot be attributed to greater spontaneous movement during the 23-h of assessment.

**Figure 4:**
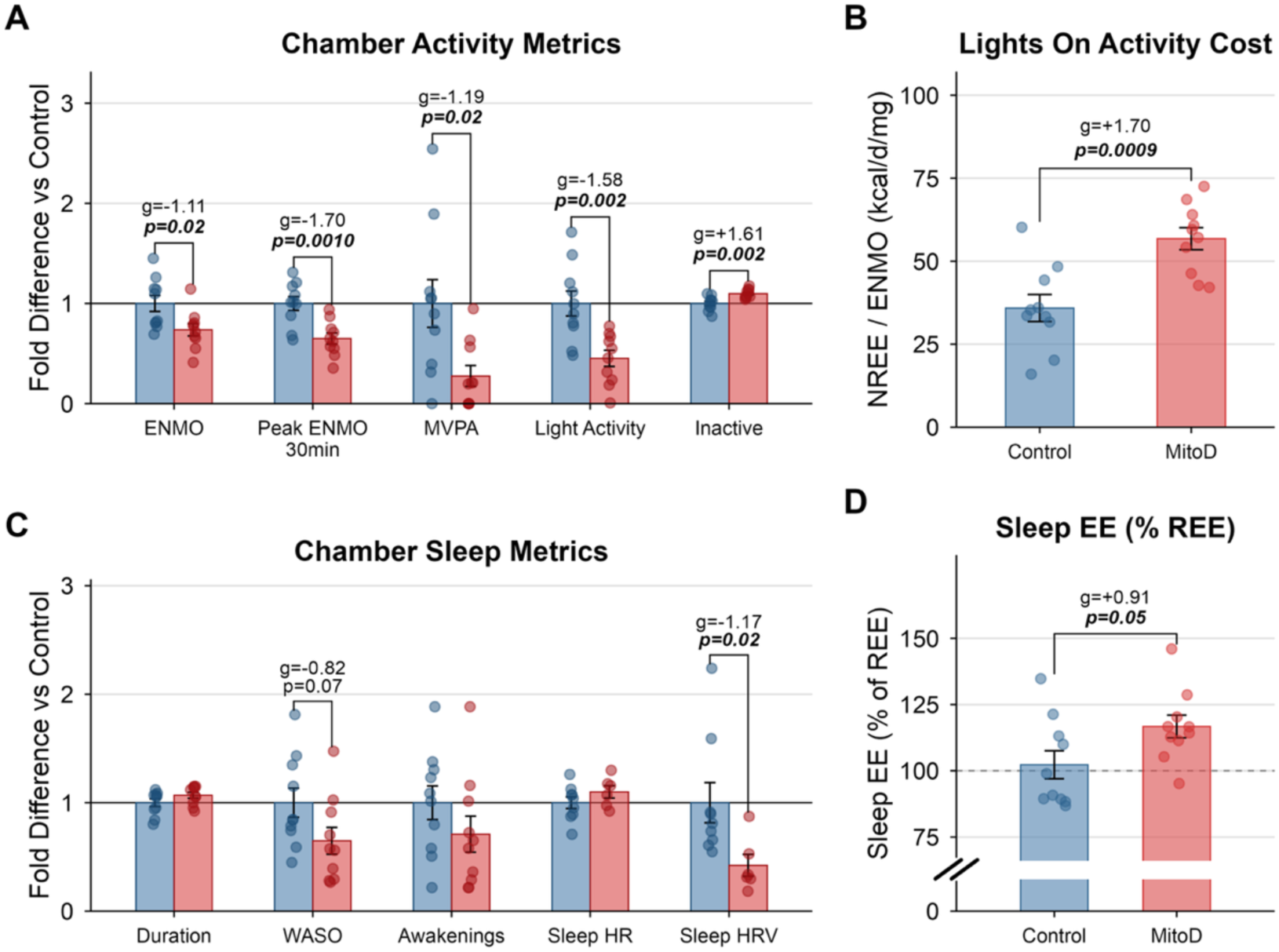
Energetics of activity and sleep in the chamber. (A) Chamber activity metrics from wrist accelerometry, expressed relative to the control mean: average ENMO, peak 30-min ENMO, moderate-to-vigorous physical activity (MVPA), light activity, and inactivity. (B) Lights-on activity cost, expressed as non-resting energy expenditure (NREE) divided by ENMO. (C) Chamber sleep metrics, expressed relative to the control mean: sleep duration, wake after sleep onset (WASO), number of awakenings, sleep heart rate, and sleep heart-rate variability (HRV). (D) Sleep energy expenditure (EE) expressed as a percentage of REE. The dashed line denotes 100% of REE. Bars show mean ± SEM with individual values. Between-group p values are from Welch’s two-sided t-tests. *Abbreviations*: EE, energy expenditure; ENMO, Euclidean Norm Minus One; HRV, heart-rate variability; MVPA, moderate-to-vigorous physical activity; NREE, non-resting energy expenditure; REE, resting energy expenditure; WASO, wake after sleep onset.

To estimate the per-unit energetic cost of activity in each group, we next examined the relationship between physical activity and non-resting energy expenditure (NREE; defined as TEE minus REE). The ratio of lights-on NREE to mean ENMO was markedly higher in MitoD than in controls (+58%; p=0.00095, **Figure 4B**), indicating greater energy expenditure per unit wrist acceleration. Normalized to unit of movement, MitoD participants expend more energy than controls.

In an exploratory analysis using 5-min bins across the lights-on WRIC period, the slope relating NREE to ENMO was also on average 48% higher in MitoD than controls, although this difference did not reach statistical significance (p=0.075, **Extended Data figure 3A**). Because wrist acceleration is not a direct measure of external work and NREE includes non-movement costs, these data do not by themselves distinguish between a higher energetic cost of movement, higher non-movement costs during waking hours, or both.

### Blunted sleep-associated metabolic suppression

Because the MitoD group displayed elevated TEE across both day and night in the WRIC (**Figure 3C**), we next examined whether differences in sleep and related physiology contributed to the nighttime EE difference. Objective sleep metrics measured by wrist actigraphy were broadly similar between groups (**Figure 4C**). Sleep duration did not differ significantly (MitoD 6.9% higher p=0.14), nor did number of awakenings (MitoD 29% lower; p=0.22), but there was a trend towards lower wake after sleep onset (WASO; MitoD 35% lower; p=0.071, **Figure 4C**). Circadian patterns in the proximal-to-distal skin temperature gradient also showed the expected day-night shift, with no significant group differences during either the day or the night (**Extended Data figure 3C**).

Despite similar sleep metrics, average EE across all sleeping minutes, expressed as a percent of REE, was significantly higher in MitoD (116.7% vs. 102.3% in controls; p=0.048, **Figure 4D**). Sleeping in the novel WRIC environment appears to have attenuated the expected sleep-associated metabolic downregulation in both groups as metabolic rate is typically 5-15% lower during sleep than in wakeful resting conditions ^53–55^. The higher-than-expected sleep EE occurred despite both groups achieving sleep efficiencies greater than 80% (Control 81.4% vs. MitoD 87.7%; p=0.08, **Extended Data figure 3D**).

The group difference in sleep EE occurred alongside differences in autonomic balance indexed by heart rate variability (HRV). During sleep, as observed in controls, HRV normally increases. Here, we found sleep HRV, calculated as root mean square of successive differences (RMSSD) to be significantly lower in MitoD (−58%; p=0.019, **Figure 4C**). Taken together, these data indicate that the blunted metabolic suppression in MitoD occurred alongside altered nocturnal autonomic physiology suggestive of tonic sympathetic nervous system activation without overt differences in sleep duration, continuity, or peripheral thermoregulation.

### Integrated 23-h WRIC patterns accompanying elevated TEE

To place the hypermetabolic 23-h WRIC phenotype in broader context, we next examined hourly group differences (**Figure 5A**) and within-subject associations with residual TEE (**Figure 5B**) across activity, metabolic, autonomic, thermoregulatory, hematologic, and mood variables.

**Figure 5:**
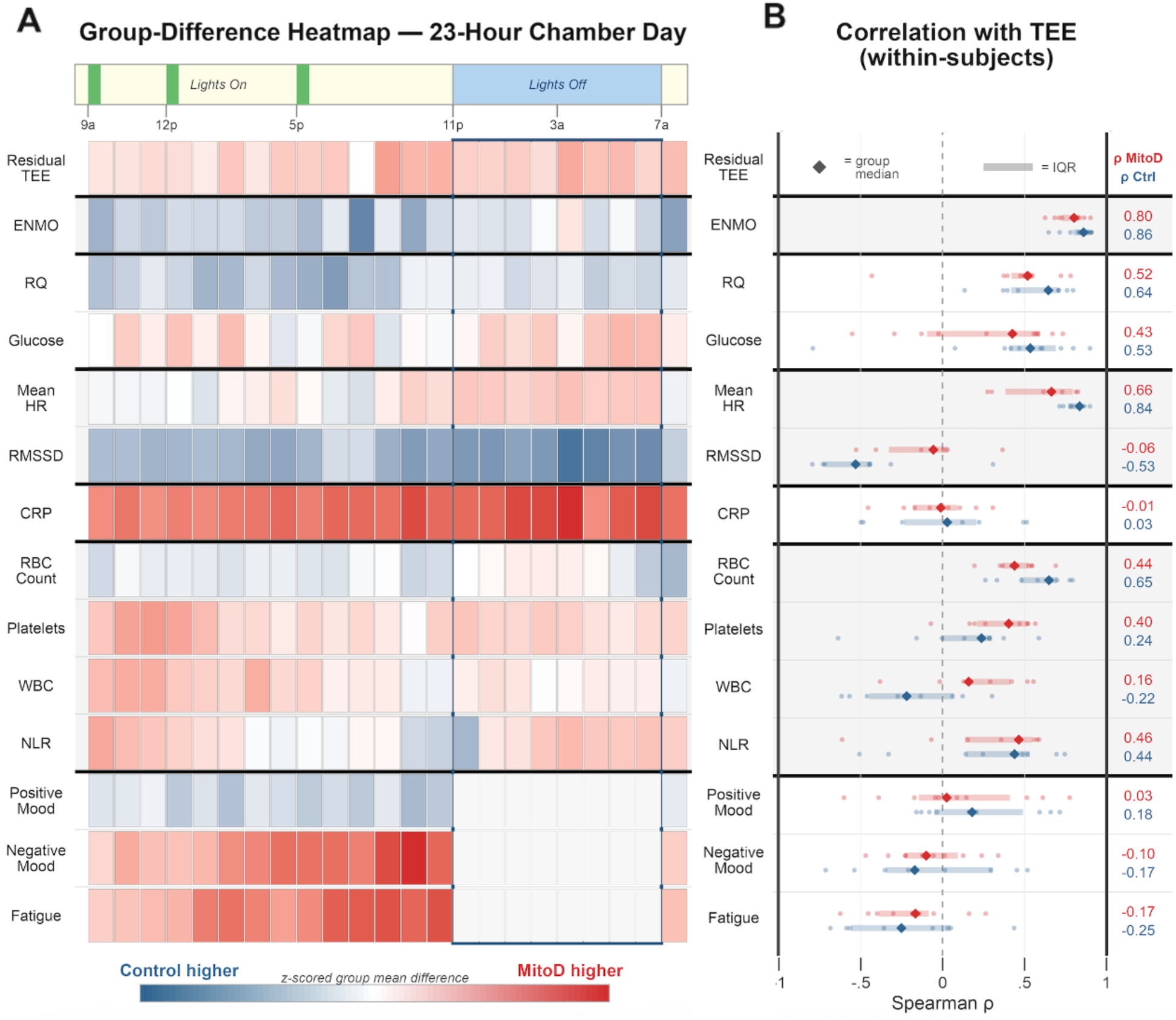
Integrated chamber day temporal patterns and correlations with TEE. (A) Group-difference heatmap across the 23-h chamber day for residual TEE and concurrently measured activity, metabolic, autonomic, inflammatory, hematologic, and mood variables. Each tile shows the z-scored group mean difference; blue indicates higher values in controls and red indicates higher values in MitoD. (B) Within-subject Spearman correlations between hourly residual TEE and each concurrently measured variable across the 23-h chamber day. Small points are subject-level correlation coefficients; diamonds and horizontal bars indicate group medians and interquartile ranges. Rightmost columns report the group-median ρ values for MitoD and controls. *Abbreviations*: CRP, C-reactive protein; ENMO, Euclidean Norm Minus One; HR, heart rate; IQR, interquartile range; NLR, neutrophil-to-lymphocyte ratio; RBC, red blood cell; RMSSD, root mean square of successive differences; RQ, respiratory quotient; TEE, total energy expenditure; WBC, white blood cell.

With respect to the temporality of group differences, four patterns emerged. First, ENMO was lower in MitoD during waking hours, but not overnight. Second, RMSSD was lower in MitoD across the 23-h WRIC day, with the largest separation during sleep, whereas heart rate was modestly higher during sleep. Third, negative mood ratings, especially fatigue, were higher in MitoD and these differences became more pronounced from morning to evening. Fourth, C-reactive protein (CRP) showed robust and persistent elevation in MitoD (+301%; p=0.0091) while hematologic variables showed comparatively modest between-group separation, consistent with a systemic inflammatory phenotype without overt immune cell alterations in MitoD (**Figure 5A; Extended Data figure 4** for individual trajectories).

Next, we performed a within-subjects analysis aimed at identifying parameters that change with energy expenditure over time (**Figure 5B**). This showed that ENMO and heart rate, as expected, were positively associated with residual TEE in both groups, indicating that TEE is higher when HR and ENMO are higher (and vice versa). RER and blood glucose showed similarly positive correlations with TEE in both groups (**Figure 5B; Extended Data figure 4**). Consistent with this, ENMO-adjusted postprandial increases in TEE did not differ between groups (**Extended Data figure 3B**), suggesting that the thermic effect of food was not altered in MitoD.

The correlation between CRP and residual TEE were near zero in both groups, consistent with the relatively slow and stable circadian kinetics of circulating CRP ^56,57^, so TEE can change from hour-to-hour independent of CRP levels (**Figure 5B**). Finally, the correlation between RMSSD and TEE was moderately negative in controls, but near zero in MitoD, consistent with a blunted nocturnal autonomic shift (**Figure 5B**).

Together, these multi-system, time-resolved, chamber-wide patterns provided the context for our next question: whether the same hypermetabolic phenotype persists under habitual free-living conditions where behavior is no longer standardized.

### Similar free-living total energy expenditure despite lower physical activity

We next aimed to determine whether the elevated TEE observed in MitoD under behaviorally clamped 23-h WRIC conditions extended to non-standardized, free-living conditions. In contrast to the WRIC findings, DLW-derived TEE averaged over 10 days did not differ between groups after adjustment for FFM (p=0.48) (**Figure 6A**). Accordingly, the chamber-to-free-living TEE increase was larger in controls (18%) than in MitoD (4.8%; p=0.049) (**Figure 6B**).

**Figure 6:**
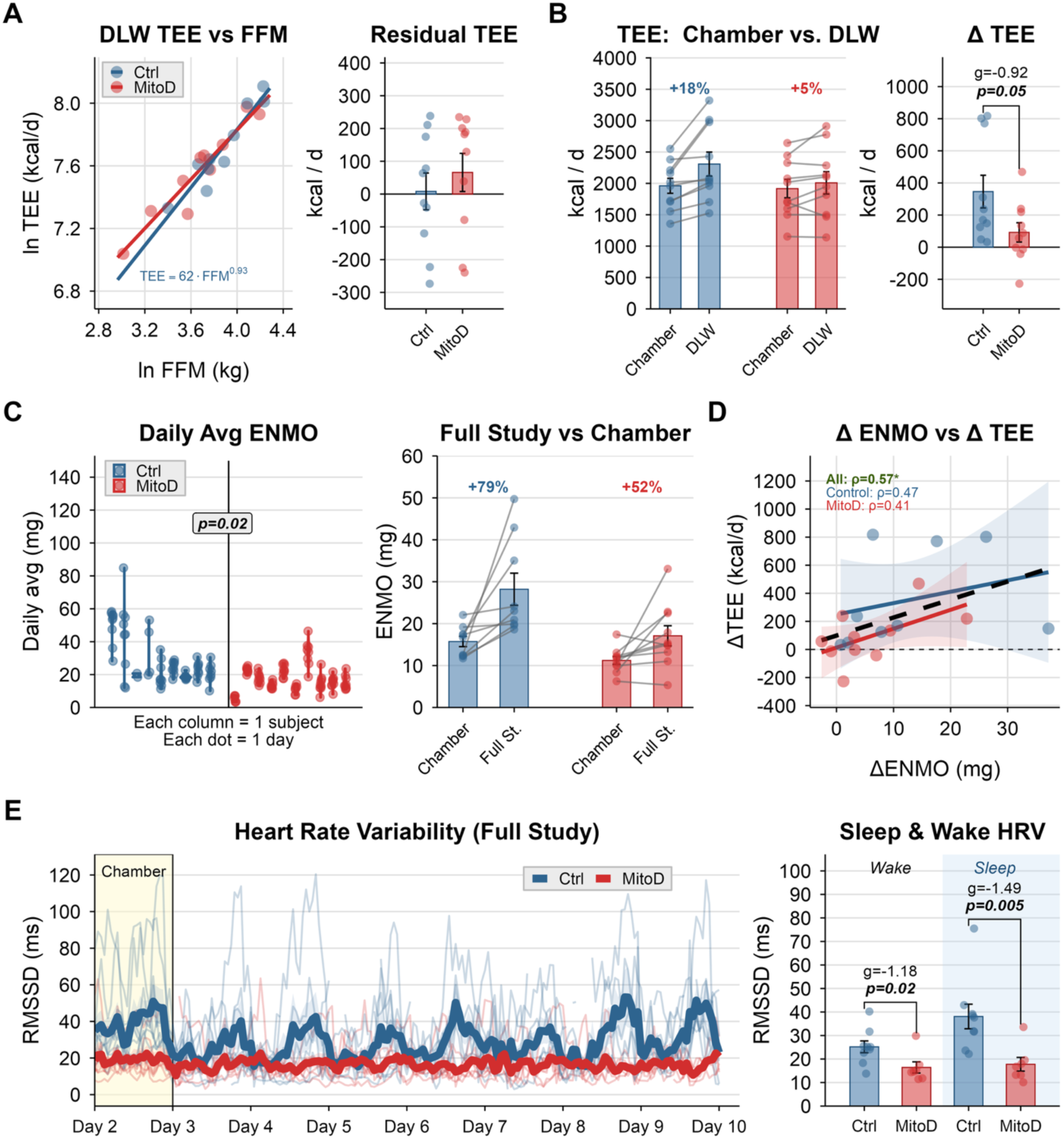
Free-living total energy expenditure, activity, and heart-rate variability. (A) Natural-log-transformed doubly labeled water (DLW)-derived total energy expenditure (TEE) plotted against natural-log-transformed fat-free mass (FFM), with adjacent FFM-adjusted residual TEE values. (B) Chamber and DLW TEE within participants, with the chamber-to-DLW difference (ΔTEE) summarized at right. (C) Daily average ENMO across post-chamber free-living days, with chamber and full-study average ENMO summarized at right. (D) Relationship between the change in ENMO from chamber to free-living conditions (ΔENMO) and the corresponding change in TEE (ΔTEE). (E) Full-study heart-rate-variability time course, with wake and sleep RMSSD summarized at right; thin lines show individuals and thick lines show group means. In panel E, the shaded region denotes the chamber study interval. In bar panels, data are mean ± SEM with individual values. Daily post-chamber activity values were compared using linear mixed-effects models with participant as a random intercept; other between-group p values are from Welch’s two-sided t-tests. In correlation panels, lines show linear fits with 95% CIs and on-panel ρ values are Spearman correlations. *Abbreviations*: DLW, doubly labeled water; ENMO, Euclidean Norm Minus One; FFM, fat-free mass; RMSSD, root mean square of successive differences; TEE, total energy expenditure.

However, the similar free-living TEE between groups occurred despite significantly lower free-living activity in MitoD. Across the free-living period, mean daily ENMO was significantly lower in MitoD than controls (−39%; p=0.022) (**Figure 6C**). Free-living sleep duration did not differ between groups (MitoD 11% higher; p=0.15) (**Extended Data figure 5A**), whereas RMSSD remained lower in MitoD during both wake (−34.8%; p=0.024) and sleep (−53.3%; p=0.0052), with the largest differences again occurring during sleep where RMSSD was not upregulated in MitoD to the same degree as in controls (**Figure 6E**). Thus, under free-living conditions, TEE and sleep are similar between groups while physical activity and HRV are significantly lower in MitoD.

Taken together, our 23-h WRIC- and free-living DLW-based measures of TEE indicated a context-specific hypermetabolic phenotype in MitoD that is evident in the WRIC but buffered in daily life. Since physical activity is the primary driver of chamber-to-free-living differences in TEE ^58^, we assessed the change in TEE and physical activity from chamber to DLW conditions (Δ = DLW − chamber). Although the chamber-to-free-living increase in ENMO was greater in controls (79%) than in MitoD (52%), the group difference was not statistically significant (p=0.19, **Figure 6C**). Nonetheless, across all participants, larger increases in average daily ENMO were associated with larger increases in TEE (Spearman ρ=0.57, p=0.012, **Figure 6D**). It follows that the context specificity of MitoD’s elevated TEE is probably related to their reduced habitual activity levels.

### Targeted inflammatory screen identifies GDF15 and CRP as biomarkers of the MitoD phenotype

GDF15 is a marker of mitochondrial stress and MitoD severity ^20,33–35^ with known effects on metabolism and behavior mediated through CNS-restricted receptors ^36^. Mean plasma GDF15 across the 23-h WRIC day, measured by ELISA, was approximately 4.6-fold higher in MitoD than in controls (1839.6 vs 404.3 pg/mL; p=0.0017, **Figure 7A**). This difference was consistent throughout the 23-h WRIC day, including the lights-on (p=0.0027) and lights-off periods (p=0.0037, **Figure 7A**). ELISA- and aptamer-based GDF15 measures were tightly correlated (Spearman ρ=0.95, p=6.2×10^−11^, **Extended Data figure 6A**).

**Figure 7:**
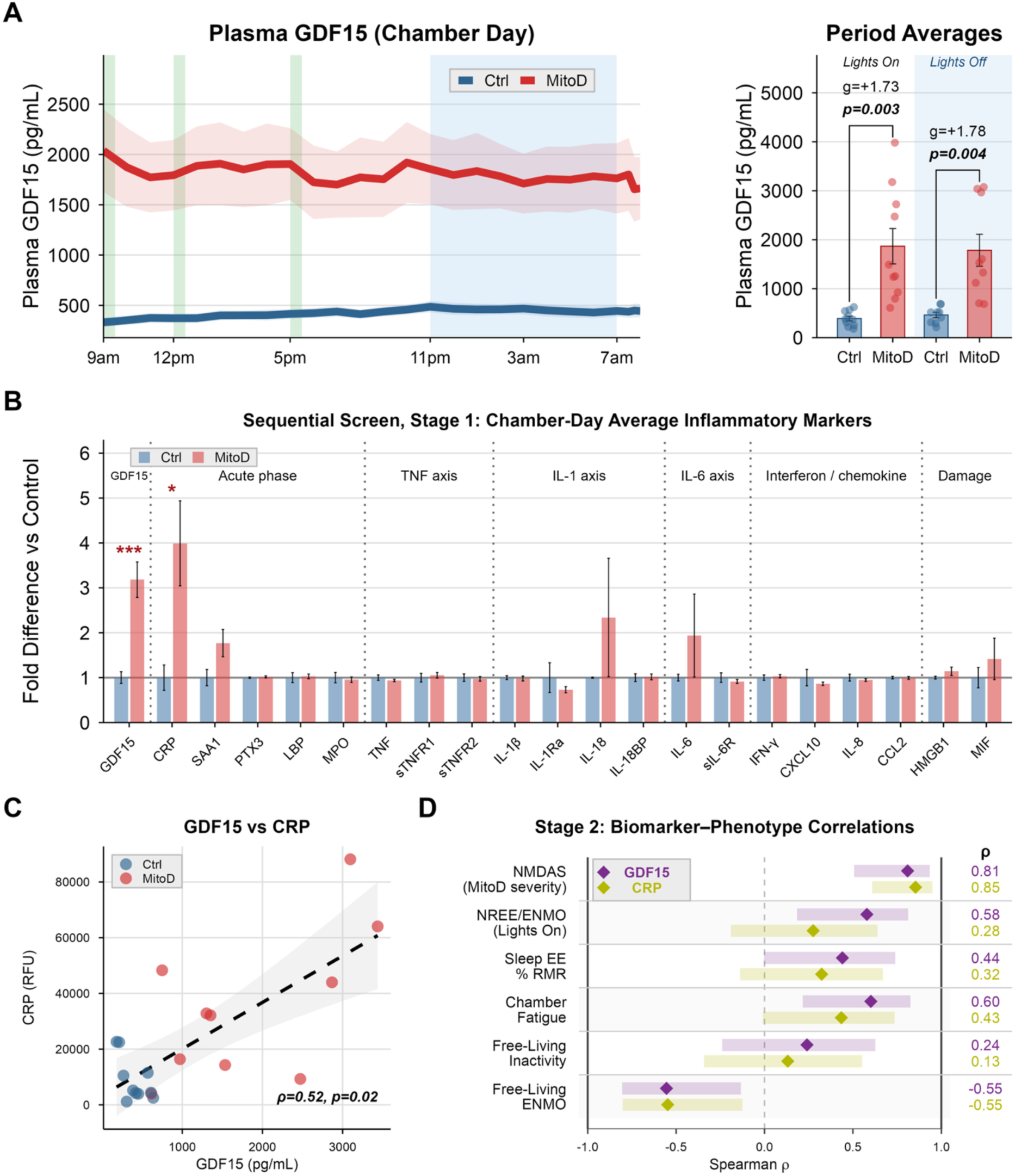
Inflammatory marker screen and biomarker-phenotype associations. (A) Chamber-day plasma GDF15 measured by ELISA, with the full time course at left (shading is SEM) and lights-on and lights-off period averages at right. (B) Stage 1 inflammatory-marker screen of 21 chamber-day average plasma markers from the SomaScan panel, expressed as fold difference relative to controls. Asterisks denote markers passing Benjamini-Hochberg-corrected one-sided Welch tests on log10-transformed chamber-day means (* q<0.05, ** q<0.01, *** q<0.001). (C) Association between subject-level mean plasma GDF15 measured by ELISA and plasma CRP measured by SomaScan. (D) Stage 2 pooled biomarker–phenotype correlations for GDF15 and CRP, the two markers passing the Stage 1 screen, across six prespecified phenotypes: mitochondrial disease severity (NMDAS), lights-on NREE/ENMO, sleep EE expressed as % REE, chamber fatigue, free-living inactivity, and free-living ENMO. In panel B, bars show mean ± SEM. In panel C, points represent individuals, the dashed line shows the pooled linear fit with 95% CI shading, and on-panel ρ and p values are pooled Spearman correlations. In panel D, diamonds and shaded rectangles indicate pooled Spearman ρ values and 95% confidence intervals; rightmost columns report the corresponding ρ values for each biomarker–phenotype pair. *Abbreviations*: CCL2, C-C motif chemokine ligand 2; CI, confidence interval; CRP, C-reactive protein; Ctrl, control; CXCL10, C-X-C motif chemokine ligand 10; EE, energy expenditure; ELISA, enzyme-linked immunosorbent assay; ENMO, Euclidean Norm Minus One; GDF15, growth differentiation factor-15; HMGB1, high-mobility group box 1; IFN-γ, interferon-gamma; IL-1β, interleukin-1 beta; IL-1Ra, interleukin-1 receptor antagonist; IL-6, interleukin-6; IL-8, interleukin-8; IL-18, interleukin-18; IL-18BP, interleukin-18-binding protein; LBP, lipopolysaccharide-binding protein; MIF, macrophage migration inhibitory factor; MitoD, mitochondrial DNA-defect group; MPO, myeloperoxidase; NMDAS, Newcastle Mitochondrial Disease Adult Scale; NREE, non-resting energy expenditure; PTX3, pentraxin 3; RFU, relative fluorescence units; REE, resting energy expenditure; SAA1, serum amyloid A1; SEM, standard error of the mean; sIL-6R, soluble interleukin-6 receptor; sTNFR1, soluble tumor necrosis factor receptor 1; sTNFR2, soluble tumor necrosis factor receptor 2; TNF, tumor necrosis factor; TEE, total energy expenditure.

Significant elevations in plasma GDF15 have been linked to reduced voluntary activity, increased fatigue, and sympathetic activation in animal ^43,44,46,47^ and human studies ^48,59–61^. Interestingly, these findings, along with results from the parent MiSBIE cohort indicating elevated markers of sympathetic outflow—urine norepinephrine (+38%, p=0.006) and blood lactate (+49%, p=0.01, **Extended Data figure 2C**)—align with the integrated MitoD behavioral and autonomic phenotype described above.

More broadly, the combination of lower free-living physical activity (**Figure 6C**), elevated CRP (**Figure 5A**), and higher fatigue (**Figure 5A**) in MitoD resembled features of inflammation-associated sickness behavior ^62^. To probe this idea, we characterized the plasma inflammatory signature that accompanied the energetic phenotype. We screened a focused panel of SomaScan 7K proteins spanning acute-phase proteins, canonical IL-1/IL-6/TNF signaling axes, interferon/chemokine mediators, damage-associated proteins, and GDF15 to identify inflammatory markers that are significantly elevated in MitoD (one-sided test). After comparing chamber day averages for each protein and correcting for multiple comparisons, only two proteins were significantly elevated in MitoD: GDF15 (q=0.00017) and CRP (q=0.018; **Figure 7B**).

We then asked which circulating marker – GDF15 or CRP – best tracked MitoD’s clinical, energetic, and behavioral phenotype. At the participant level, ELISA-measured plasma GDF15 and SomaScan CRP were moderately correlated across the full sample (Spearman ρ=0.52, p=0.019, **Figure 7C**), indicating that the two markers captured partially overlapping but non-identical information. Because both biomarkers differed markedly by group, pooled correlations are best interpreted as associations across the case-control gradient rather than as evidence of within-group effects (see **Extended Data figure 6B**). Within that framework, GDF15 and CRP tracked equally well with higher mitochondrial disease severity (NMDAS) and lower free-living activity (**Figure 7D**). However, GDF15 consistently showed moderately stronger associations with the energetic and subjective components of the phenotype, including higher waking NREE/ENMO, chamber fatigue, and sleep EE (**Figure 7D**), lending partial support for a body-to-brain signaling axis linking OxPhos defects to participant behavior and experiences.

## DISCUSSION

We report that under behaviorally clamped WRIC conditions, adults with pathogenic mtDNA defects exhibited higher FFM-adjusted TEE across both day and night, despite lower spontaneous activity and normal sleep metrics. By contrast, over the 10-day free-living period, DLW-based TEE was similar between groups, despite significantly lower physical activity in MitoD. Together, these findings demonstrate a persistent energetic burden in people with mitochondrial OxPhos defects, exposed under standardized WRIC conditions but masked by lower physical activity under free-living conditions. We confirm that plasma GDF15 is markedly elevated in MitoD and demonstrate that GDF15 tracks with CRP, higher perceived fatigue, lower activity, and higher 23-h WRIC energetic burden. Thus, our data identifies GDF15 as a biomarker of a fatigue-related phenotype, consistent with other studies ^43,59,60,63^.

Methodologically, the new insights reported here emerged from the combination of three contrasting EE measurement contexts: i) 1-h REE measurement that controls for activity, posture, feeding, stress, and sleep, ii) WRIC-based TEE assessment where physical activity, meal energy and nutrient composition, and sleep are highly standardized, and iii) 10-d DLW protocol when constraints on behavior are removed.

The resulting findings suggest that relying only on standard REE assessments, DLW-based TEE and/or the physical activity level ratio (PAL; TEE/REE) may systematically underestimate disease-associated energy costs. In a previous study of MELAS patients, REE and accelerometer-derived PAL were used to estimate energy requirements, leading to the conclusion that TEE is lower in MitoD than in controls ^24^. Our findings bring this conclusion into question and suggest that the relationship between objectively measured physical activity and the PAL ratio is disrupted in MitoD, rendering this parameter inadequate. Supporting this notion, our data shows the PAL ratio to similar between groups despite significantly lower habitual activity levels in MitoD (**Extended Data figure 5D-E**). This discordance likely reflects some combination of movement inefficiency and non-movement energy costs, neither of which are captured by the 1-h REE measurement used in our or the previous study. Thus, WRIC-based TEE measures, particularly when coupled with time-resolved measures of activity, sleep, and other psychobiological processes, are well suited to detect otherwise hidden (i.e., behaviorally buffered) differences in energy expenditure and allocation. This appears as a meaningful lesson to guide future research in human energy expenditure.

Three findings from the WRIC period support the existence of waking hypermetabolism in MitoD: i) the elevated chamber energy expenditure despite lower physical activity, ii) the elevated ratio of NREE to mean wrist acceleration (NREE/ENMO), and iii) the trend toward steeper NREE-ENMO slopes. These converging findings are consistent with higher movement-associated energy costs in MitoD. Because wrist ENMO is an imperfect proxy for external work and NREE includes non-movement costs, these data cannot distinguish the contribution of muscle contraction from other non-movement costs. For example, systemic inflammation (of either immune or non-immune origin) indexed by elevated CRP, could reflect a non-movement contributor to the NREE/ENMO finding in MitoD. Nevertheless, our interpretation leans on exercise studies showing elevated oxygen consumption per unit work ^22,64^, together with hyperkinetic circulatory responses with impaired peripheral oxygen extraction in MitoD ^21,65,66^. This evidence of inefficiency suggests the mitochondria-associated hypermetabolism uncovered here involves at least some contribution of chemomechanical inefficiency.

A strength of this study is the continuous and combined analysis of metabolic rate, objective sleep metrics and autonomic physiology during sleep. Two novel findings are noteworthy. First, MitoD displayed elevated sleep EE relative to controls despite similar sleep duration, wake after sleep onset, and number of awakenings. Second, sleep HRV was significantly lower in MitoD, suggesting autonomic involvement in the sleeping hypermetabolism phenotype. Together, these findings show that OxPhos defect-related hypermetabolism isn’t confined to waking, active periods, but also extends to sleep—when physical activity is near zero in both groups. The blunted sleep-associated metabolic suppression in MitoD may partially explain the profound fatigue and related functional impairments that MitoD patients experience ^17^.

In contrast to the WRIC findings, FFM-adjusted free-living TEE measured over the full study period by DLW suggested the existence of compensatory behavioral mechanisms to prevent chronic hypermetabolism. One plausible interpretation is that the higher disease-associated costs observed under WRIC conditions limit the fraction of the energy budget available for discretionary activity (i.e., people feel less vigorous), consistent with the constrained model of TEE ^28,32,67^. Energetic tradeoffs are known to occur in the context of acute infection ^62^, childhood immune burden ^68,69^, pregnancy ^70^, and aging ^71^ but to our knowledge, they have not been reported in chronic genetic diseases. Whether fatigue and physical inactivity in mitochondrial disorders represent an adaptive attempt to balance the energy budget, or are merely consequences of disease burden, remains an open question.

If bioenergetic inefficiency or other hypermetabolism-driving processes were causally related to behavioral phenotypes of physical inactivity and subjective fatigue, we would expect body-to-brain signals to inform the central nervous system of the organism’s energetic state—a form of interoception specific to the systemic metabolic state, or “metaboception.” The GDF15-GFRAL axis is one such candidate mechanism. GDF15 is produced by peripheral tissues ^33^ and acts through its brainstem-restricted receptor GFRAL^38–40^ to regulate visceral malaise states ^41,42^, food intake ^42^, physical activity ^43,44^ and autonomic physiology ^37,46,47^ across a broad range of conditions ^36^. As expected ^20,33–35^, GDF15 was persistently elevated across the chamber day in MitoD. Moreover, higher GDF15 tracked with disease severity, CRP, greater fatigability, higher chamber fatigue ratings, higher NREE/ENMO, higher sleep EE, and lower free-living activity, as would be expected if GDF15 contribute to produce these energy-conservation states. Human experimental studies support causal relevance of GDF15-GFRAL signaling in other disease contexts ^44^, and genetic studies implicate fetal GDF15 production in hyperemesis gravidarum ^78^. Establishing a causal link between GDF15 and features of human mitochondrial diseases will require intervention studies targeting the GDF15-GFRAL axis, although these should be undertaken with caution ^73^.

Finally, a central unresolved question in mitochondrial medicine is how OxPhos defects produce multisystem disease and reduced lifespan ^16^. One concept that may bridge the psychobiology of OxPhos defects, hypermetabolism, and lifespan is *allostatic load*—the cumulative wear and tear on physiological systems from sustained activation of energetically costly stress-responsive systems ^4,75,73,76,77^. In our MitoD cohort and others, multiple findings are consistent with elevated allostatic load in MitoD. This includes the blunted sleep-related metabolic suppression, low HRV, systemic inflammation, resting plasma lactate ^20^, elevated urinary norepinephrine ^4^, and persistent fatigue ^17^. Outside of MitoD, chronic psychosocial stress activates many of the same physiological systems that are dysregulated in our MitoD cohort ^78^. Therefore, we speculate that the energetic burden associated with stress responses, which we recently discovered to acutely induce GDF15 ^33^, could help explain why chronic stress generally promotes broad, multisystem pathology and may thus contribute to early mortality.

In conclusion, we have combined state-of-the-art EE measurement methods with continuous measurements of physical activity, sleep, autonomic function, mood, plasma inflammatory markers, and GDF15 to examine the effects of mtDNA defects on human energy expenditure. We find that mitochondrial OxPhos defects impose energy costs that are concealed under tightly controlled REE measurement conditions, revealed under standardized WRIC conditions, and buffered in daily life by behavioral compensation. GDF15 emerges as a potential metaboceptive signal that could mediate such compensation, a hypothesis to be validated in further studies. These findings call for studies using WRIC and DLW in combination to uncover potentially hidden, context-dependent hypermetabolism across disease and health conditions.

### Limitations of the study

This study has several limitations. First, the cohort was small (n=10 per group) and genetically heterogeneous, which precluded subtype analyses. Technically, the movement-cost inference is limited by the use of wrist accelerometry and by the fact that NREE includes non-movement costs. Moreover, the nighttime WRIC findings were influenced by the novel sleeping environment of the room calorimeter while participants were wearing a number of unfamiliar sensor devices including an indwelling intravenous line used for hourly blood draws. The lower daytime RER may reflect altered substrate handling, negative energy balance, or both. Finally, the cross-sectional design and the pooled nature of the GDF15 associations do not resolve whether lower activity reflects compensation for higher costs, direct symptom limitation, GDF15-mediated effects, or some combination. Resolving these questions in future will yield exciting new insights around the timing and regulation of energy human expenditure and allocation.

## Acknowledgements

We are grateful to the parent MiSBIE study team (www.picardlab.org/MiSBIE) and participants who made this study possible, to the Bionutrition Research Core (UL1TR001873), the Irving Institute Clinical Research Resource staff (UL1TR001873), the Center for Advanced Laboratory Medicine (CALM), and the members of the Mitochondrial Psychobiology laboratory at CUIMC.

## Author contributions

M.P., A.S., M.H., D.G., M.-P.S.-O., M.R., S.A.C. and E.L.M. designed the study. E.D.S. and A.S. led study operations. S.R., and S.L. coordinated study visits. K.W. oversaw metabolic chamber operations and calorimetry data processing. F.M.Z. analyzed sleep and ASA24 data. H.S. designed the chamber diets and analyzed chamber food intake. Q.H., A.J., D.K. and F.M.Z supported chamber-day operations and biospecimen collection and processing. M.K. and D.S. performed GDF15 assays. J.C. processed ECG data. D.G. designed and oversaw the body composition measurements. W.S. quantified MRI-derived organ and fat volumes. N.B.A. contributed parent MiSBIE data and analyses used in Extended Data figure 2. C.K. and K.E. supported participant recruitment. H.P. and C.T. contributed to study strategy and interpretation of results. S.A.C. and E.L.M. performed doubly labeled water analyses. M.H. and M.R. supervised the clinical portion of the study. E.D.S. analyzed the data and drafted the manuscript. M.P. supervised the project and edited the manuscript. All authors reviewed, revised, and approved the final version of the manuscript.

## Declaration of Generative AI and AI-Assisted Technologies in the Writing Process

During preparation of this manuscript, the authors used generative AI tools, including ChatGPT (OpenAI) and Claude (Anthropic), to improve readability, refine phrasing, summarize background literature for author review, suggest potentially relevant references, and provide feedback on clarity and organization. The authors independently evaluated all AI-assisted suggestions, verified cited literature against original sources, and revised the manuscript using their own scientific judgment. The authors take full responsibility for the content of the manuscript.

## EXTENDED DATA FIGURES

**Extended Data figure 1:**
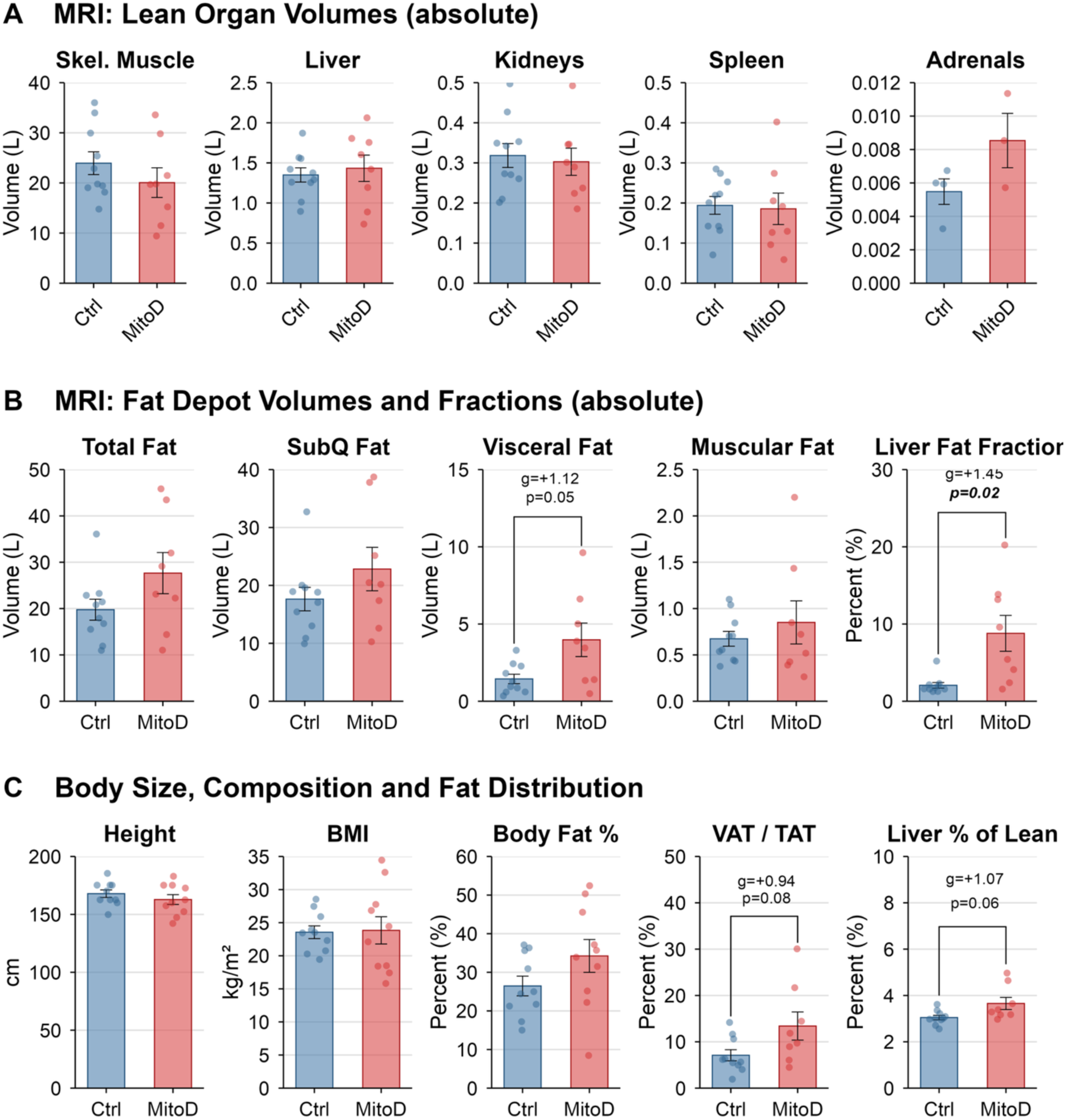
Body composition, organ volumes, and fat distribution values. (A) Absolute MRI-derived volumes of skeletal muscle, liver, kidneys, spleen, and adrenal glands. (B) Absolute MRI-derived volumes of total adipose tissue, subcutaneous adipose tissue, visceral adipose tissue, and intramuscular adipose tissue, plus liver fat fraction. (C) Height, body mass index (BMI), body fat percentage, visceral adipose tissue as a fraction of total adipose tissue (VAT/TAT), and liver volume as a percentage of total lean volume. Bars show mean ± SEM with individual values. Between-group p values are from Welch’s two-sided t-tests and are shown where indicated on the panels. *Abbreviations*: BMI, body mass index; MitoD, mitochondrial DNA-defect group; MRI, magnetic resonance imaging; TAT, total adipose tissue; VAT, visceral adipose tissue.

**Extended Data figure 2:**
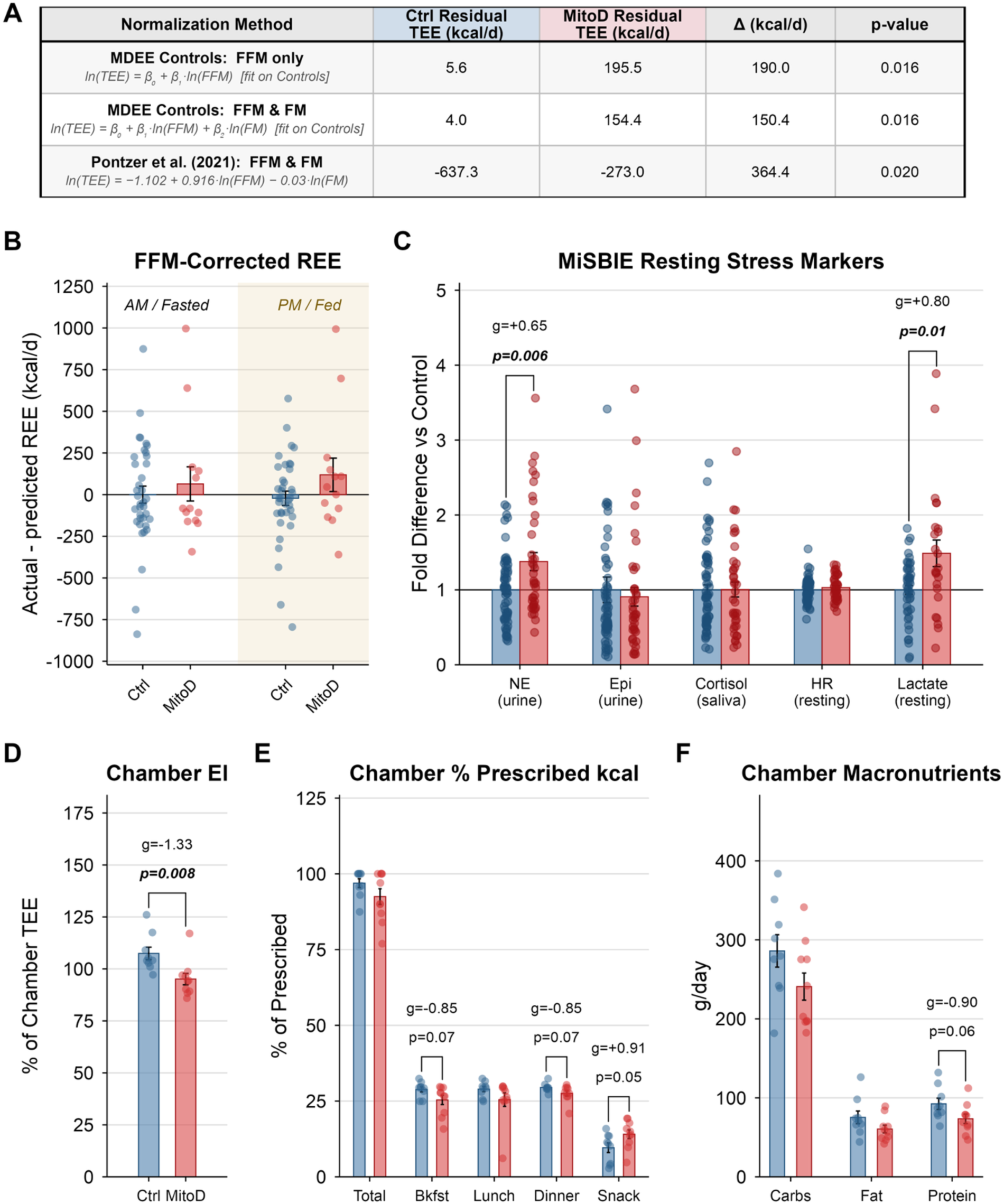
Alternative chamber TEE normalization, MiSBIE resting energy expenditure, and MDEE chamber dietary intake. (A) Comparison of group differences in residual chamber TEE across three normalization methods: two allometric models fit on MDEE Control subjects [ln(TEE) ∼ ln(FFM) and ln(TEE) ∼ ln(FFM) + ln(FM)] and a published allometric equation from the IAEA Doubly Labeled Water Database (n=6,421; adults aged 20–60 years). The table reports mean residual TEE per group, the MitoD–Control difference (Δ), and Welch’s t-test p value for each method. (B) Fat-free-mass (FFM)-adjusted resting energy expenditure (REE) measured in the larger parent MiSBIE cohort under AM/fasted and PM/fed conditions. (C) MiSBIE resting stress-marker fold-difference chart showing urinary norepinephrine (NE), urinary epinephrine (Epi), salivary cortisol, resting heart rate (HR), and resting lactate, each expressed relative to the control mean. (D) Chamber energy intake (EI) expressed as a percentage of chamber TEE. (E) Actual chamber intake expressed as a percentage of prescribed kilocalories, with total intake and meal-specific intake at breakfast, lunch, dinner, and snack. (F) Chamber macronutrient intake. Panel A uses MDEE chamber and body-composition data. Panels B–C use data from the larger parent MiSBIE cohort. Panels D–F use chamber data from the MDEE study. In panel B, shaded halves denote the AM/fasted and PM/fed conditions. Bars show mean ± SEM with individual values. Between-group p values are from Welch’s two-sided t-tests and are shown where indicated on the panels. *Abbreviations*: EI, energy intake; Epi, epinephrine; FFM, fat-free mass; FM, fat mass; HR, heart rate; MiSBIE, Mitochondria Stress, Brain Imaging, and Epigenetics study; NE, norepinephrine; REE, resting energy expenditure; TEE, total energy expenditure.

**Extended Data figure 3:**
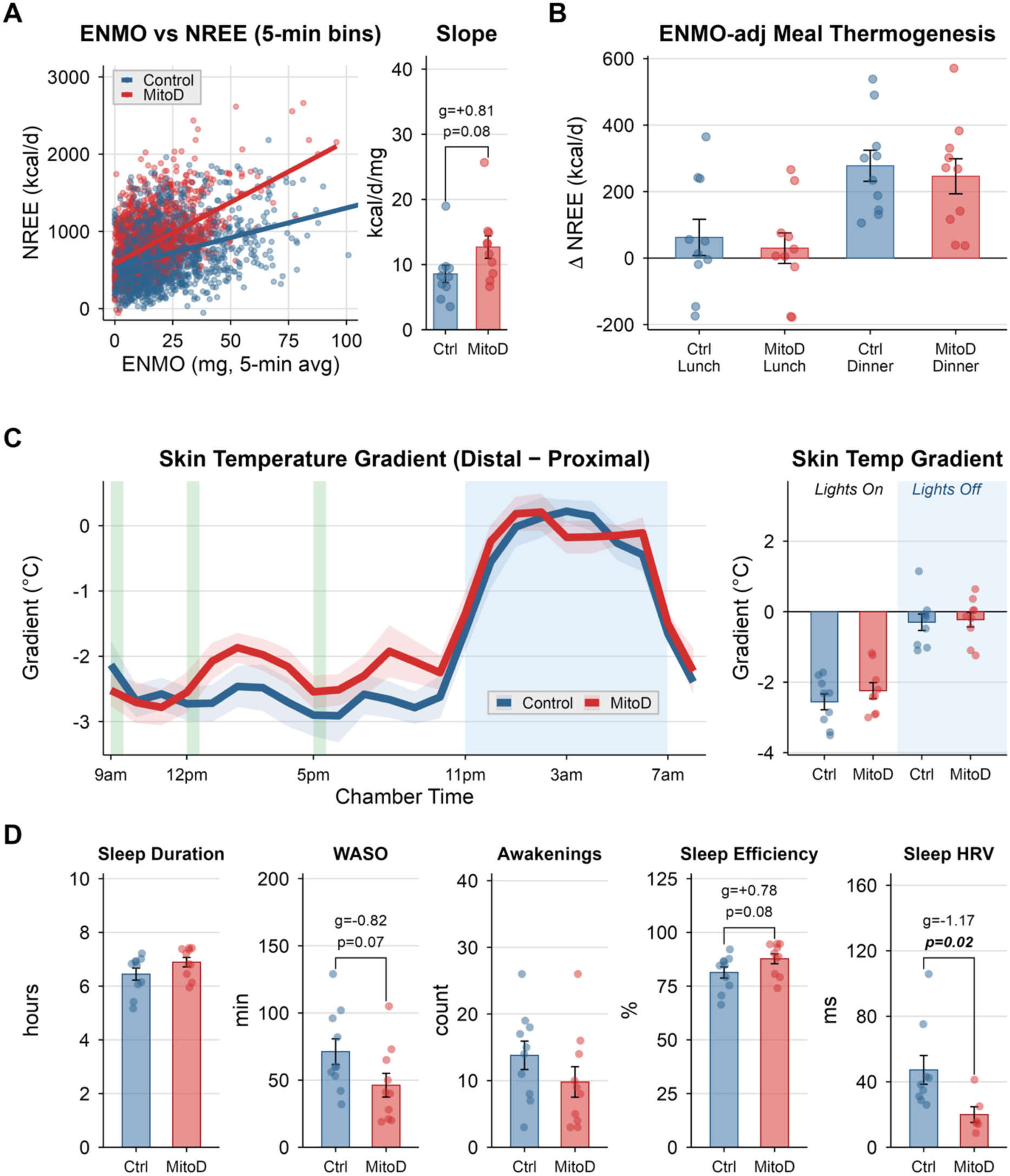
Movement cost, meal thermogenesis, circadian skin temperature gradient, and chamber-night sleep metrics. (A) Relationship between chamber non-resting energy expenditure (NREE) and ENMO across 5-min bins, with subject-level NREE/ENMO slopes summarized at right. (B) ENMO-adjusted change in NREE around lunch and dinner. (C) Distal-proximal skin temperature gradient across the chamber day, with lights-on and lights-off summaries at right. (D) Chamber-night sleep metrics shown as actual subject-level values: total sleep duration, wake after sleep onset (WASO), number of awakenings, sleep efficiency, and sleep heart rate variability (HRV; RMSSD during lights-off). Green vertical bands indicate standardized meals and blue shading indicates the lights-off interval. In panel A, lines show linear fits. In panel C, lines and shading indicate mean ± SEM. Bar panels show mean ± SEM with individual values. Between-group p values are from Welch’s two-sided t-tests where indicated. Abbreviations: ENMO, Euclidean Norm Minus One; HRV, heart rate variability; NREE, non-resting energy expenditure; RMSSD, root mean square of successive differences; WASO, wake after sleep onset.

**Extended Data figure 4:**
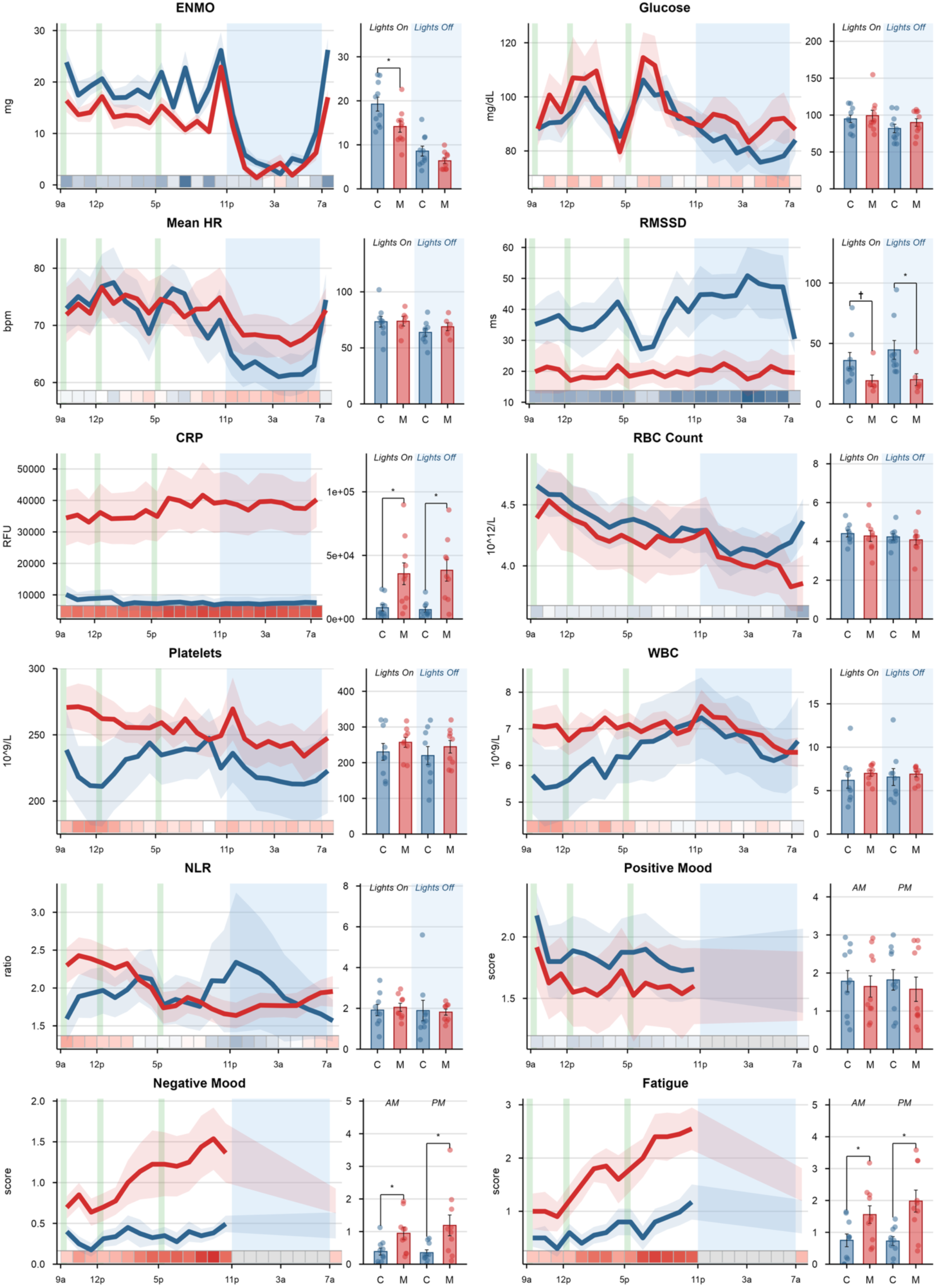
Quantitative chamber-day trajectories for variables highlighted in Figure 5. Time-course plots show chamber-day group means ± SEM for ENMO, glucose, mean heart rate, RMSSD, CRP, red blood cell count, platelet count, white blood cell count, neutrophil-to-lymphocyte ratio, positive mood, negative mood, and fatigue. Companion bar plots summarize lights-on and lights-off means for physiologic variables and AM versus PM means for mood variables, with individual values overlaid. The thin bottom ribbon in each time-course panel reproduces the z-scored group-difference heatmap from Figure 5A for the same variable. Green vertical bands indicate standardized meals and blue shading indicates the lights-off interval. Symbols above the bar plots indicate between-group period-comparison statistics where shown on the panels. *Abbreviations:* C, control; CRP, C-reactive protein; ENMO, Euclidean Norm Minus One; HR, heart rate; M, mitochondrial DNA-defect group; NLR, neutrophil-to-lymphocyte ratio; RBC, red blood cell count; RFU, relative fluorescence units; RMSSD, root mean square of successive differences; WBC, white blood cell count.

**Extended Data figure 5:**
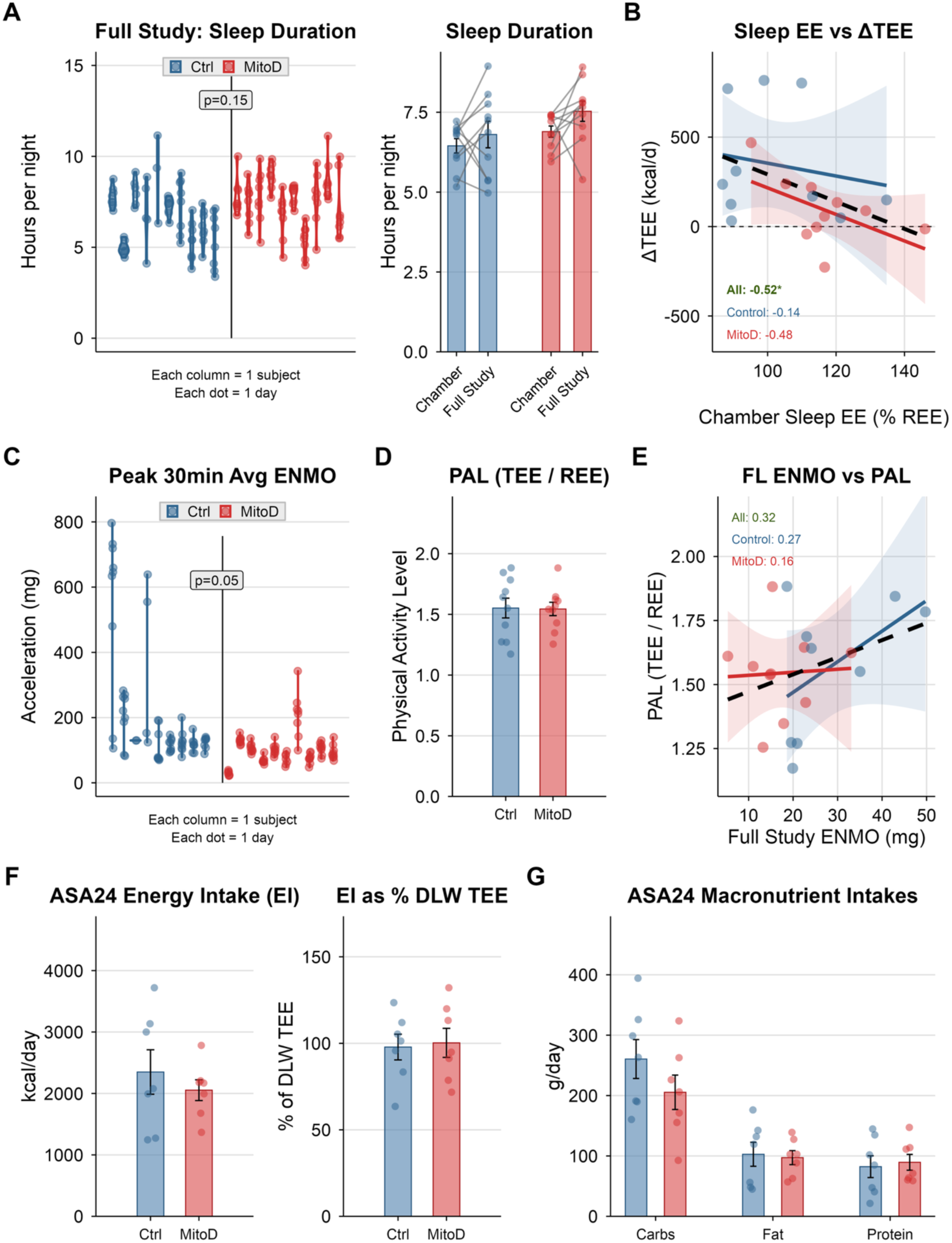
Free-living sleep duration, peak activity, physical activity level, and dietary intake. (A) Nightly sleep duration across post-chamber free-living days, with chamber and full-study sleep duration summarized at right. (B) Relationship between chamber sleep EE (% REE) and the chamber-to-DLW change in TEE (ΔTEE). (C) Peak 30-min ENMO across post-chamber free-living days. (D) Physical activity level (PAL), defined as DLW-derived TEE divided by REE, compared between groups. (E) Relationship between full-study average ENMO and PAL. (F) ASA24-derived daily energy intake and energy intake expressed as a percentage of DLW-derived TEE. (G) ASA24-derived macronutrient intakes. In panels A and C, each column represents one participant, each dot represents one day. Bar panels show mean ± SEM with individual values. Daily free-living sleep duration and peak ENMO were compared using linear mixed-effects models with participant as a random intercept. PAL and dietary intake were compared using Welch two-sample t-tests. In correlation panels, lines show linear fits with 95% CIs and on-panel ρ values are Spearman correlations. *Abbreviations*: ASA24, Automated Self-Administered 24-h Dietary Assessment Tool; DLW, doubly labeled water; EE, energy expenditure; ENMO, Euclidean Norm Minus One; PAL, physical activity level; REE, resting energy expenditure; TEE, total energy expenditure.

**Extended Data figure 6:**
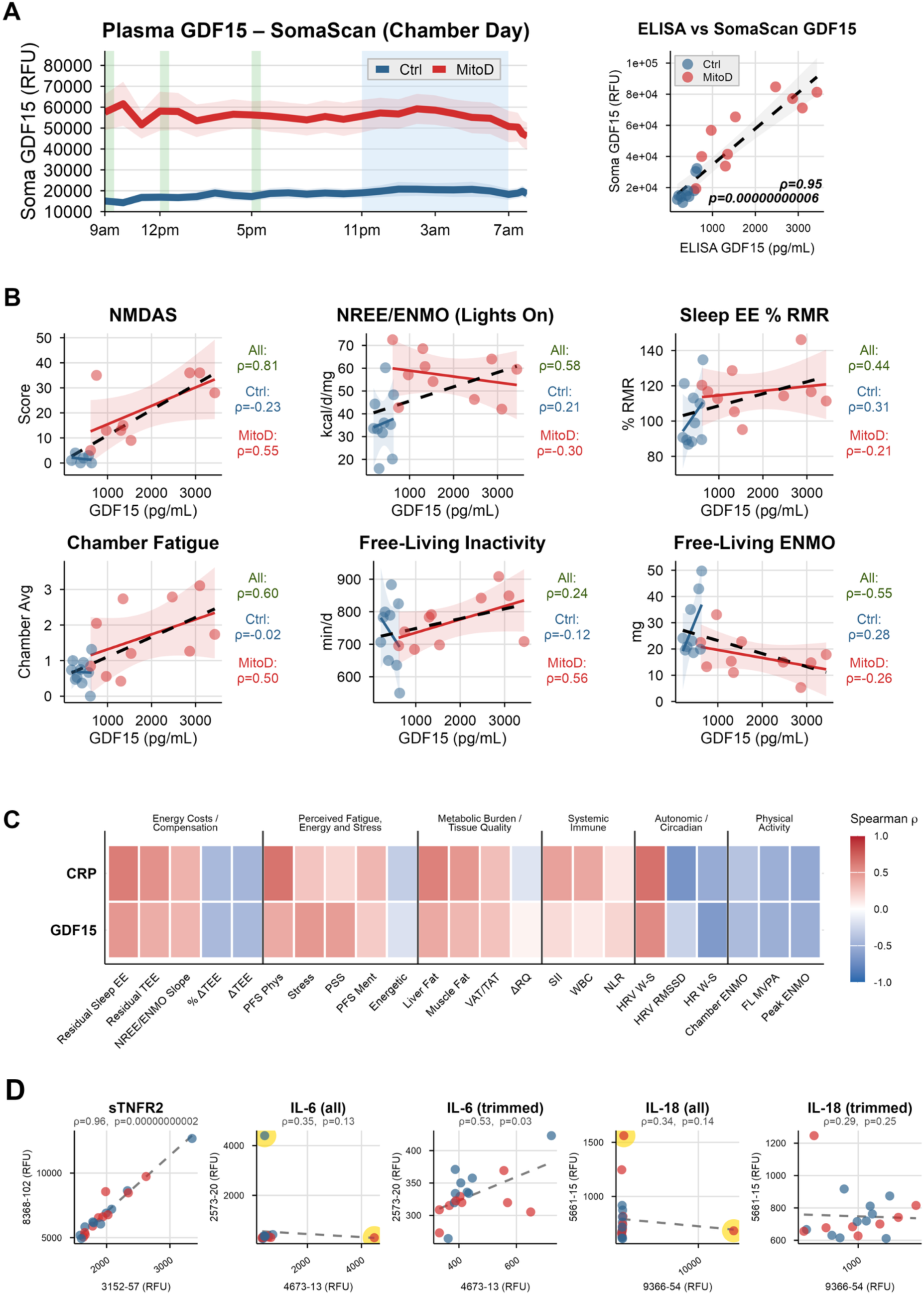
Plasma GDF15 characterization, phenotype correlations, assay concordance, and extended biomarker-phenotype associations. (A) Chamber-day plasma GDF15 measured by SomaScan, with the full timecourse at left (shading is SEM) and the cross-platform association between ELISA and SomaScan chamber-day means at right. All time points for each participant are averaged into a single value to estimate the most stable estimate of GDF15 for each person. (B) Scatter plots of chamber-day mean plasma GDF15 (ELISA) versus each of the six phenotype outcomes from the Figure 7D forest plot, shown separately by group. Row 1: NMDAS, NREE/ENMO (Lights On), Sleep EE % REE. Row 2: Chamber Fatigue, Free-Living Inactivity, Free-Living ENMO. (C) Heatmap of pooled Spearman correlations for CRP and GDF15 across additional phenotype variables not shown in Figure 7D, spanning energetic burden/compensation, perceived fatigue/energy/stress, metabolic burden/tissue quality, systemic immune, autonomic/circadian, and physical activity domains. (D) Duplicate SOMAmer concordance plots for sTNFR2 and for paired IL-6 and IL-18 aptamers, shown for all subjects and, for IL-6 and IL-18, after trimming outlier subjects flagged [yellow highlight] by a 3×IQR rule on either aptamer. Axis labels identify the paired SOMAmer sequence identifiers. In panel A, lines show group means, green vertical bands indicate standardized meals and blue shading indicates the lights-off interval in the time-course panels. In panel A right and panel D, points represent individuals and dashed lines show linear fits; on-panel ρ and p values are Spearman correlations. In panel B, points represent individuals, solid colored lines show group-specific linear fits with 95% confidence bands, and dashed black lines show pooled linear fits; side panels report Spearman ρ for all subjects, controls, and MitoD separately. In panel C, cells are colored by pooled Spearman ρ, with blue indicating negative correlations and red indicating positive correlations. *Abbreviations*: CRP, C-reactive protein; EE, energy expenditure; ELISA, enzyme-linked immunosorbent assay; ENMO, Euclidean Norm Minus One; FL, free-living; GDF15, growth differentiation factor-15; HR, heart rate; HRV, heart-rate variability; MVPA, moderate-to-vigorous physical activity; NLR, neutrophil-to-lymphocyte ratio; NMDAS, Newcastle Mitochondrial Disease Adult Scale; NREE, non-resting energy expenditure; PFS, Pittsburgh Fatigability Scale; PSS, Perceived Stress Scale; RFU, relative fluorescence units; REE, resting energy expenditure; RMSSD, root mean square of successive differences; RQ, respiratory quotient; SII, systemic immune-inflammation index; SOMAmer, Slow Off-rate Modified Aptamer; TEE, total energy expenditure; VAT/TAT, visceral adipose tissue as a fraction of total adipose tissue; W-S, wake minus sleep; Δ, change.

## METHODS

### EXPERIMENTAL MODEL AND STUDY PARTICIPANT DETAILS

#### Human participants

Adults aged 18–60 years were recruited into two groups: healthy controls and individuals with genetically confirmed pathogenic mitochondrial DNA (mtDNA) defects. The mitochondrial disease group included participants with either the m.3243A>G mtDNA point mutation or single large-scale mtDNA deletions. The final MDEE cohort included 10 individuals with mtDNA defects and 10 healthy controls.

Participants with mitochondrial disease were recruited through established clinical and research pathways, including referral by co-investigator Dr. Michio Hirano from his clinical program and from the NIH m.3243A>G natural-history cohort. Initial outreach was performed by a certified genetic counselor or research coordinator before referral to the study team for full screening. Healthy controls were recruited through RecruitMe, Columbia University Irving Medical Center’s IRB-approved volunteer registry.

All study procedures were conducted at Columbia University Irving Medical Center under Institutional Review Board protocol AAAT5410. Written informed consent was obtained electronically through REDCap before any research procedures were performed. Final eligibility was confirmed on Study Day 1 by physical examination and, where applicable, urine pregnancy testing.

#### Screening and eligibility

Screening was conducted by structured telephone or secure-Zoom interview and included medical history, medication use, sleep patterns, and lifestyle factors. Exclusion criteria included pregnancy, shift work, sleep disorders, current malignancy or chronic inflammatory disease, MRI-incompatible implants or severe claustrophobia, tobacco or other substance use, and excessive alcohol consumption, defined as ≥14 drinks per week in women or ≥21 drinks per week in men.

#### Cohort characterization

Participant demographics, anthropometrics, and body composition characteristics are reported in ***Figure 2A***. Demographic variables included age, sex, race/ethnicity, employment status, height, body mass index, and body fat percentage. The mitochondrial disease cohort included participants with genetically confirmed mtDNA defects, comprising five individuals with the m.3243A>G point mutation and five individuals with single large-scale mtDNA deletions.

Because of the small sample size and genetic heterogeneity of the mitochondrial disease group, analyses were performed using a combined mitochondrial DNA-defect group rather than stratifying by mtDNA variant class.

#### Parent MiSBIE cohort

Some supplemental analyses used data from the larger parent Mitochondrial Stress, Brain Imaging, and Epigenetics study. These parent-cohort analyses were limited to the MiSBIE-derived measures shown in ***Extended Data figure 2***, including fasted morning and fed afternoon resting energy expenditure, urinary norepinephrine and epinephrine, salivary cortisol, resting heart rate, and resting lactate. For these supplemental analyses, individuals with pathogenic mtDNA defects were analyzed as a combined mitochondrial disease group. Details of the MiSBIE study design have been previously described, and the specific MiSBIE methods used for the present analyses are provided under Method details.

### METHOD DETAILS

#### Mitochondrial Daily Energy Expenditure (MDEE) Study Design

This study employed a two-phase protocol consisting of an intensive 2.5-day in-person visit to the medical center (in-patient for the 23-h whole room indirect calorimeter [WRIC] period), followed by 8.5 days of at-home monitoring (see **Figure 1**).

Prior to Day 1, participants stayed at a nearby hotel and began fasting in the evening >12h prior to arriving at the medical center. On Day 1, participants underwent a 60-min resting energy expenditure (REE) assessment in a WRIC (7:30-8:30am) and had their body-composition analyzed by quantitative magnetic resonance (QMR; 8:45-9:15am). Participants then completed a urine void and consumed their personalized dose of doubly labeled water (DLW; ∼9:20am) followed by a whole-body MRI to measure organ volumes and fat distribution (∼11am). Subsequently, participants were fitted with a wrist-worn accelerometer (Actigraph GT3X), Holter style ECG device (MOX3, Maastrichts Instruments) and continuous glucose monitor (CGM; FreeStyle Libre Pro) (∼1pm). Urine samples were collected 4- and 6-hours post-DLW dose (∼1:20pm and ∼3:20pm). Between these timepoints, participants consumed a standardized snack and were evaluated by a physician. Between study days 1 and 2, participants returned to the hotel and began fasting before 9pm.

On Day 2, participants were admitted to the inpatient clinical research unit and had their vitals assessed before entering the metabolic chamber at 9am for a 23-h stay. While in the room calorimeter, participants followed a standardized meal schedule, consumed isocaloric and nutrient-matched meals designed by the CUIMC Bionutrition Research Core. Participants also completed a questionnaire package including hourly mood assessments. At 11pm, participants were instructed by study staff to turn off the lights, silence all devices and attempt to initiate sleep. Participants were given a wake up call by study staff at 7am the next morning; wake up was verified by lights on and/or text message response. Participants exited the room calorimeter just after 8:00am on Day 3. While in the room calorimeter, blood was drawn via a ∼8-feet intravenous line fed through an airtight port, every hour, on the hour (and more frequently upon awakening), as shown in **Figure 1**.

During the free-living phase (Days 3–11) participants were instructed to continue wearing the wrist-based accelerometer and ECG devices at all times and to fill out daily sleep diaries each morning upon awakening. In addition, participants were instructed to collect their morning urine (second void) each day of the free-living period and were assigned three web-based ASA24 dietary recalls, to be completed on non-consecutive days encompassing two weekdays and one weekend day. DLW kinetics from clinic and home urine provided total energy expenditure (TEE), while WRIC data yielded resting, sleeping, and activity-derived components, integrating confined/controlled and free-living/habitual energetics within a single protocol.

#### REE by 1-h WRIC

WRIC was used to measure morning fasted REE on Day 1 (separately from the 23-h WRIC TEE measurement that occurred on Days 2-3). WRIC has the highest precision and accuracy for REE measurement of all available methodologies ^79^. The room calorimeter at CUIMC is comprised of an air-tight, temperature-controlled room connected to gas sensors and flow meters. By pulling a known amount of fresh air through the room, respiratory gases from the study participant are sampled on the exhaust side of the system for measurement of O2 and CO2 using fuel cell oxygen and near infrared carbon dioxide sensors (Model GA-3m2, Sable Systems Intl, Las Vegas, NV). Analysis software for the SW-Promethion System is installed on a Sable Systems approved computer, with a Sable Systems Promethion Interface Module (Model IM-2). Data are recorded and processed by the Sable software programs Caloscreen and Expedata. Rates of energy expenditure (EE) were calculated using the Weir equation ^80^ to convert the steady state concentrations of respiratory gases (VO_2_ L/min and VCO_2_ L/min) to EE in kilocalories per min (kcal/min). The coefficient of variation for REE VO2 and VCO2 for quality control during the period of data collection was 2.78 and 2.14 respectively.

Participants were instructed to fast for at least 12-h prior to the REE measurement (7:30pm the night before). On the morning of the measurement (Day 1), participants were transported by car from a nearby hotel to the medical center and were brought to the calorimetry facility by wheelchair. Upon arrival at the calorimetry facility, participants were instructed to leave all electronic devices outside the chamber and to lay still in a supine position on the bed with head slightly elevated on a pillow for the duration of the measurement (1-h). Lights were kept dimmed and the ambient temperature was kept between 23-25C. Study staff monitored participants to ensure they remained still and awake. The first and last ten minutes of data were discarded to allow for the room and sample line air to reach steady state and eliminate anticipatory reactions from the participant that may artificially inflate EE. For each subject, all contiguous 10-min windows in the cleaned recording were evaluated, and REE was defined as the window with the lowest mean EE among windows with a within-window coefficient of variation ≤5%.

#### Anthropometric and body composition measurements

Anthropometric and body composition assessments were conducted on Day 1 in the Body Composition Unit of the New York Nutrition Obesity Research Center at CUIMC immediately after the REE measurement described above. Participants, still in the fasted state, were instructed to empty their bladders, remove clothing and jewelry, and wear a provided hospital gown and slippers immediately before anthropometric measurements. Urine was collected as a baseline sample for the DLW analysis (see below). Body weight was measured to the nearest 0.1 kilogram (kg) using a calibrated scale (Ohaus Champ General Purpose Scale, Ohaus Corp., Pine Brook, NJ, USA), while height was measured to the nearest 1 millimeter (mm) using a wall-mounted stadiometer (Holtain Ltd., Crymych, UK).

Quantitative magnetic resonance (QMR; EchoMRI^TM^ Adult Human Body Composition Analyzer, Echo Medical Systems, Houston TX, USA) is a noninvasive technique that employs proton nuclear magnetic resonance to measure body composition. The system can accommodate individuals up to 250 kg and is standardized to detect changes in fat mass (FM) with a precision (replicated measurements CV) of < 0.5%. Duplicate measurements were performed by a trained technologist, lasting 6 minutes in total. The system provides estimates of FM, lean mass, free water, and total body water (TBW). We analyzed FM and fat-free mass (FFM), which was calculated by subtracting FM from measured body weight.

n=2 participants in the MitoD group had metal implants that were incompatible with QMR-based body composition measurement. These participants had their body composition measured by bioelectrical impedance analysis using the Tanita-305 device (Tanita Corp., Tokyo, Japan) ^81^.

#### Organ volumes and fat distribution by whole-body MRI

Whole-body MRI was performed on a 3.0T General Electric system (SIGNA Premier, Milwaukee, WI). Participants were positioned in a supine position with their arms stretched overhead in the scanner. The scanner was landmarked at the L4-L5 intervertebral disc, with 1 cm-slice images taken from L4-L5 to the top of the fingerprints and from L4-L5 to the end of the toes. Image slices were spaced 4 cm apart. The organ protocol included contiguous slices of 0.4 cm in thickness from the dome of the liver to the bottom of the kidneys. Liver fat was evaluated with Iterative Decomposition of water and fat with Echo Asymmetry and Least Square Estimation (IDEAL-IQ) Fat Fraction sequence ^82^. Imaging analysis was performed to quantify volumes (L) for the whole body, total and compartmental adipose tissue, and skeletal muscle. Data were acquired and analyzed as described previously ^83,84^.

#### Free-living TEE by DLW

TEE was measured over the full study period using DLW. Prior to dosing, a baseline urine sample was obtained for determination of background enrichments of ^2^H and ^18^O. The DLW dose was determined based on TBW (estimated as 73% of FFM, measured by QMR) to provide 2.2g ^18^O (10% APE) and 0.15g ^2^H (99.8% APE) per kg TBW ^85^. Participants consumed the DLW followed by two, 100g rinses of bottled water immediately after QMR on Day 1. To capture peak isotope enrichment in participants’ body water, post-dosing urine samples were obtained 4-h (PD4) and 6-h (PD6) after DLW dosing. Further urine samples were collected each morning of the free-living period (second void of the day). Baseline, PD4, PD6, day 7 and day 10 urine samples were shipped on dry ice from CUIMC to the CU Anschutz DLW Isotope Core for analysis.

Frozen urine samples were thawed, prepared by centrifugation and analyzed for ^18^O and ^2^H enrichment by Off-Axis Integrated Cavity Output Spectroscopy (OA-ICOS, ABB Inc., Zurich, Switzerland), as previously described ^86^. Briefly, OA-ICOS uses a laser-based methodology and poses advantages of increased throughput and thus faster analysis of samples as compared to traditional isotope ratio mass spectrometry (IRMS). We have recently reported ^87,88^ that TEE (i.e. CO_2_ production) measured using OA-ICOS over 7-d is valid compared to simultaneous whole-room indirect calorimetry and IRMS. Data were analyzed using commercially available Post Analysis Software (LWIA Post Analysis, ABB Inc., Version 4.4.1.1), which utilizes inter-run standard measurements to automatically calibrate isotope measurements. Samples were analyzed in duplicate, and repeated if the SD exceeded 26 for ^2^H and 0.56 for ^18^O. Dilution spaces for ^2^H and ^18^O were calculated from the baseline samples according to the methods of Coward ^89^. Total body water was calculated as the average of dilution spaces of ^2^H and ^18^O after correction for isotopic exchange with other body pools ^90^. CO_2_ production rate was calculated using the two-point method and the recently published equation of Speakman et al. ^91^. TEE was be calculated using the Weir equation ^80^, assuming a respiratory quotient of 0.86, and averaged over 10-d. The average within subject CV for repeat measures of TEE in the CU Anshutz DLW Isotope Core is 6.1±3.8%.

#### TEE by 23-h WRIC

Participants were instructed to fast for at least 12-h before the start of their 23-h stay in the room calorimeter (9:00pm Day 1). On the morning of the measurement (day 2), participants were transported by car from a nearby hotel to the medical center (to avoid potential confounding effects of physical activity), were admitted to the hospital as in-patient research participants and brought to the calorimetry facility by wheelchair. The room calorimeter is equipped with a toilet, sink, cot, television and windows facing outside and into the control room. The 23-h measurement period began at 9:00am and concluded the following morning just after 8:00am. During this time, participants followed a standard schedule consisting of three meals at pre-determined times (9am, 12pm and 5pm), a snack at a time of the subject’s choosing (recorded by study staff) and lights-off for sleep between 11pm-7am. All foods were prepackaged and taken into the metabolic chamber upon participant entry. There were hourly blood draws and mood surveys with more frequent draws and surveys between 7am-8am (7am, 7:30am, 7:45am, 8am) to capture the awakening response.

All food consumed in the room calorimeter was prepared and provided by the CUIMC Bionutrition Research Core. Energy requirements were estimated using the Mifflin-St Jeor equation with an individualized activity factor. Diets were standardized to provide 50% carbohydrate, 18% protein, and 32% fat, with daily energy distributed across three meals (30% each) and one snack (10%). Menus were developed and analyzed using the Nutrition Data System for Research (NDSR), version 2018 (Nutrition Coordinating Center, University of Minnesota, Minneapolis, MN), to closely match target energy and macronutrient distributions. Mealtimes were fixed and consumption was directly observed by study staff. Participants were instructed to consume all foods provided. Any uneaten food was collected and weighed, and nutrient analysis was performed via NDSR to determine actual intake. Two participants received individualized diets for religious practices or food allergies, matched as closely as possible to prescribed energy and macronutrient targets. Water was provided ad libitum; no non-study beverages were permitted.

#### Actigraphy-based sleep assessment using ActiLife software

To measure sleep and physical activity across the full study period, participants were fitted with a tri-axial accelerometer with ambient light sensor (ActiGraph GT3X+, Ametris [formerly ActiGraph Corp.], Pensacola, FL), hereafter referred to as an Actigraph. They were instructed to wear the Actigraph at all times (24-h/day) on their non-dominant wrist for the entire study period except for during water-based activities (i.e. swimming, bathing, etc.) and to maintain a log listing exact times at which the device was removed and reapplied.

For sleep analysis, actigraphy was supplemented by sleep diaries that participants completed each morning upon awakening; diaries included information on the time participants got into bed, attempted to initiate sleep, overnight awakenings, and the time(s) of morning awakenings. Nocturnal sleep was quantified using the ActiLife software from Ametris. The validated Cole-Kripke ^92^ and Tudor-Locke ^93^ algorithms were applied to raw accelerometer data for determination of sleep vs. wake epochs and sleep period detection, respectively. Information from the daily sleep diaries, along with visual inspection of the data, were used to enhance the accuracy of bed- and wake-time estimates determined from the algorithms. Actigraphy-derived sleep outcomes of interest included participant nightly averages for total sleep time (TST), sleep efficiency (% time in bed spent asleep), wake after sleep onset (WASO, hours), and number of awakenings.

Primary sleep outcomes were derived from expert-scored ActiLife files rather than from GGIR summary outputs described below. Nights failing prespecified quality-control criteria, including non-wear, zero counts despite reported wear, excessive time in bed, or implausible summary values, were excluded from aggregation. GGIR-derived sleep/wake labels were used only for secondary time-aligned physiologic summaries, including sleep- and wake-specific HRV estimates.

#### Actigraphy-based physical activity assessment using GGIR

Raw accelerometer files were processed in R using GGIR version 3.2.6. Processing was based on raw wrist acceleration with Euclidean Norm Minus One (ENMO, mg) used as the primary activity metric. The GGIR workflow used the default GGIR autocalibration settings, imputation of short non-wear periods, advanced-format sleep logs, and exported both summary outputs and 60-second time-series files for downstream integrations. Activity intensity was categorized using ENMO thresholds of <40 mg for inactivity, 40 to <100 mg for light activity, 100 to <400 mg for moderate activity, and >=400 mg for vigorous activity; moderate-to-vigorous physical activity (MVPA) was defined as the sum of moderate and vigorous activity minutes.

For the 23-h WRIC day activity analyses, accelerometry was processed using a subject-specific activity-log file supplied through the q-window setting to isolate the chamber interval. Because the 23-h WRIC stay crossed midnight, the WRIC period was represented by two GGIR segments spanning WRIC entry to midnight and midnight to WRIC exit. WRIC-day mean ENMO was calculated as the duration-weighted mean across these two segments. WRIC-day inactivity, light activity, moderate activity, vigorous activity, and moderate-to-vigorous physical activity (MVPA) were calculated by summing the corresponding segment-level durations across the two WRIC segments. WRIC-day minute-level accelerometry used in downstream time-aligned analyses was obtained from the exported 60-second GGIR part 5 time-series files. To preserve both halves of the 23-h WRIC stay across midnight, the WRIC-segment GGIR setup used relaxed part 5 validity settings, including segment wear validity of 25%, no sleep-period requirement for segment retention, and a minimum part 5-d criterion of 6-h.

For free-living activity analyses, the same GGIR workflow generated midnight-to-midnight daily summaries across the full recording period. In this setup, days were defined using GGIR’s MM time window with a day border of 00:00. Participant sleep diaries were provided to GGIR as advanced sleep logs with sleep-window type set to sleep period time. Additional day-level exclusions were applied to days with inadequate quality, including GGIR cleaning code ≥2, >25% non-wear, or missing ENMO values. Free-living activity outcomes were calculated as participant-level means across valid post-chamber days.

Although GGIR processing also generated sleep-related outputs, these were not used for the primary sleep outcomes reported in this manuscript. Instead, all reported sleep outcomes were derived from semi-manual expert scoring in ActiLife by an experienced scorer using diary-guided review and visual inspection (described above), an approach endorsed by the American Academy of Sleep Medicine and Sleep Research Society for free-living sleep assessment ^94^. GGIR-derived sleep/wake labels were used only when a minute-level sleep-versus-wake classification was needed for secondary derived physiologic measures, such as sleep and wake HRV summaries.

#### Free-Living Dietary Intakes

Self-reported dietary intakes were assessed via multiple 24-h dietary recalls using the Automated Self-Administered 24-h® (ASA24®) Dietary Assessment Tool developed by the National Cancer Institute ^95^. Participants were instructed to use this web-based tool to record all foods and beverages consumed over 3 non-consecutive 24-h days; reporting days were assigned to participants and comprised at least 2 weekdays and one weekend day over the 8-day at home period. Participants were given written instructions on how to use the tool as well as links to video instructions and an online ‘demo’ diary for practice. In addition, participants were encouraged to take photos of each eating/drinking occasion in real time to assist in completing the corresponding recall and enhance accuracy of reporting. Participants were requested to complete an additional 24-h recall if they initially submitted incomplete or extreme entries (≤ 2 recalls or 24-h energy intake ≤ 500 kcal). Nutrient and energy intakes were computed automatically by ASA24 using the USDA Food and Nutrient Database. For analysis, recall days were excluded when ASA24 flagged the report as extreme or when reported intake was <800 kcal/day, and nutrient intakes were averaged across remaining valid recalls for each participant.

#### Blood Collection and Processing

During the 23-h stay in the room calorimeter, hourly blood samples were obtained through an indwelling venous catheter in either the AC joint or forearm connected to extension tubing passed through an airtight port, allowing repeated sampling without entering the chamber or disturbing the participant. The catheter was flushed with saline, secured to the arm, and maintained with a continuous infusion of 0.9% normal. A total of 26 blood draws were targeted: hourly on the hour from 09:00am on Day 2 through 07:00am on Day 3, followed by additional post-awakening draws at 07:30am, 07:45am, and 08:00am. At each draw, one 1ml EDTA-treated tube (BD; purple top) was collected for complete blood count (CBC) analysis and two 2.5ml citrate-treated tubes (BD; teal/blue top) were collected for plasma-based assays, including GDF15. Total planned blood volume was approximately 156 mL across the chamber stay.

Purple-top tubes for CBC were refrigerated immediately and transferred to Columbia’s Center for Advanced Laboratory Medicine (CALM) laboratory for analysis within 12-h of collection. Teal-top tubes were centrifuged at 1000 × g for 5 min, after which plasma was aspirated and transferred to a 15-mL conical tube. The recovered plasma was then centrifuged again at 2500 × g for 20 min, aliquoted into 1.5-mL tubes, and stored frozen at -80C until analysis.

#### Complete Blood Count

Complete blood counts (CBC) with differential were measured by the Center for Advanced Laboratory Medicine (CALM) lab at CUIMC. Assays yielded standard hematologic measures, including hemoglobin, hematocrit, white blood cell count, red blood cell count, platelet count, red blood cell indices, and leukocyte differential measures with absolute counts. From these data, CBC-derived inflammatory indices were calculated, including the neutrophil-to-lymphocyte ratio (NLR; absolute neutrophil count / absolute lymphocyte count). CBC measures were aligned to protocol timepoint and used to generate chamber-day hourly trajectories and participant-level summary measures; missing or quality-control-flagged draws were excluded from aggregated analyses.

Blood collections were flagged as diluted (saline contamination from the IV catheter flush) when at least 2 concentration sensitive measures (Hemoglobin, Hematocrit, RBC) dropped more than 15% from the previous consecutive draw while both MCV and MCH changed less than 5%, confirming the drop reflects dilution rather than a true physiological change (since dilution lowers absolute counts but leaves red cell indices stable). Blood collections flagged as diluted (n=12 of 499 successful blood collections, 2.4%) were removed from the GDF15 and SomaScan inflammatory marker analyses described below.

#### Plasma GDF15

Plasma GDF15 concentration was quantified using a high-sensitivity ELISA kit (R&D Systems, DGD150, SGD150) per manufacturer instructions. Plasma samples were diluted 1:4 for these assays. Final plates were read by absorbance at 450 nm on a SpectraMax M2 (SpectraMax Pro 6, Molecular Devices), and concentrations were interpolated using a four-parameter logistic (4PL) model in GraphPad Prism (version 10.3.1) using a standard curve run on each plate. Plasma reference samples were run on each plate, and their inter-assay coefficients of variation (CVs) were monitored. All samples from the time course for a particular individual were kept within the same assay batch. Samples were run in duplicate and final concentrations were calculated as the average of these duplicates. If the CV between duplicates was larger than 15%, the sample was rerun when sufficient sample remained. If rerunning was not possible, the sample was omitted from downstream analyses. Samples corresponding to blood collections flagged for saline contamination (e.g. dilution; see Complete Blood Count, above) were excluded from the analyses.

#### Circulating Inflammatory Markers by SomaScan Proteomics

Circulating inflammatory markers were measured in citrate plasma samples collected during the 23-h chamber stay using the SomaScan v4.1 proteomic assay (SomaLogic Operating Co., Boulder, CO). The SomaScan platform uses modified single-stranded DNA aptamers (SOMAmers) to quantify protein concentrations for approximately 7,000 analytes simultaneously, with protein abundance reported in relative fluorescence units (RFU). Hourly aliquots of citrate plasma were shipped on dry ice to SomaLogic for assay. Sample processing, hybridization normalization, median signal normalization, plate scaling, calibration, and adaptive normalization by maximum likelihood (ANML) were performed by SomaLogic according to standard operating procedures. Data were returned as a single .adat file with sample-level (RowCheck) and analyte-level (ColCheck) quality control flags.

From the full SomaScan panel, we selected 21 primary markers representing an a priori inflammation panel: C-reactive protein (CRP), lipopolysaccharide-binding protein (LBP), myeloperoxidase (MPO), pentraxin 3 (PTX3), serum amyloid A1 (SAA1), high mobility group box 1 (HMGB1), macrophage migration inhibitory factor (MIF), interleukin-18 (IL-18), IL-18 binding protein (IL-18BP), IL-1 receptor antagonist (IL-1Ra), IL-1β, IL-6, soluble IL-6 receptor (sIL-6R), monocyte chemoattractant protein-1 (CCL2/MCP-1), interferon gamma-induced protein 10 (CXCL10/IP-10), interferon-γ (IFN-γ), IL-8 (CXCL8), tumor necrosis factor-α (TNF-α), soluble TNF receptor I (sTNFR1/TNFRSF1A), and soluble TNF receptor II (sTNFR2/TNFRSF1B). Growth differentiation factor-15 (GDF15) was included as a 21st marker to enable cross-platform comparison with the ELISA-based GDF15 measurements. Three proteins (IL-6, sTNFR2, IL-18) were each targeted by two independent SOMAmers on the SomaScan panel; for these, both the primary and secondary aptamers were extracted to assess within-platform concordance. Duplicate SOMAmer reagents were highly concordant for sTNFR2 but less so for IL-6 and IL-18, indicating that duplicate aptamers for some targets should not be treated as interchangeable without caution (**Extended Data figure 6D**).

Data were parsed from the .adat file using the SomaDataIO R package. For the primary SomaScan screen, a draw-by-analyte observation was considered clean only when it passed sample-level SomaLogic QC, had a finite RFU value, was not linked to a CBC-defined diluted blood draw, and was not an outlier by the within-participant 3-MAD rule. Analyte-level ColCheck flags were retained for reporting but were not used as an exclusion criterion in the primary screening analysis. For each participant, the chamber-day mean RFU for each of the 21 primary markers was computed as the arithmetic mean of clean draws across all available timepoints.

For the CRP and GDF15 SomaScan timecourses (**Figure 5A**, **Figure 7A** and **Extended Data figure 6A**), clean draw-level RFU values were retained at each canonical timepoint and summarized by chamber period (lights-on, lights-off, awakening). The duplicate SOMAmer concordance analysis (**Extended Data figure 6D**) included both primary and secondary aptamers for IL-6, sTNFR2, and IL-18, with ColCheck relaxed for this comparison so that both aptamers were available regardless of QC flag status. Per-subject chamber means (mean=20.1, median=22, range=3-26 hourly timepoints per subject) for each aptamer were compared by Spearman correlation and Bland-Altman analysis to assess within-platform measurement agreement.

To identify circulating inflammatory markers that differed between groups and to examine their associations with the energetic and behavioral phenotype, we implemented a two-stage sequential screen. In Stage 1, chamber-day mean RFU values for each of the 21 protein markers were compared between MitoD and controls using one-sided Welch’s t-tests on log10-transformed values (alternative hypothesis: MitoD > Control). P values were adjusted for multiple comparisons across the 21 markers using the Benjamini-Hochberg procedure, and markers with adjusted q < 0.05 were advanced to Stage 2. Individual subject values were plotted as fold-difference relative to the control group mean. In Stage 2, markers that passed the screen, together with plasma GDF15 (ELISA), were correlated with six prespecified phenotype variables — mitochondrial disease severity (NMDAS total score), chamber NREE/ENMO, sleep EE as a percentage of REE, average chamber fatigue rating, free-living inactivity, and free-living mean ENMO — using pooled Spearman rank correlations. Hedges’ g was computed as the standardized effect size for all Stage 1 between-group contrasts.

#### ECG-based Heart Rate and HRV

Participants were fitted on day 1 with a chest-worn ambulatory ECG device (MOX3; Maastricht Instruments) and were instructed to wear the device at all times (24-h/day) for the entire study period except during water-based activities (ie. swimming, bathing, etc.) and to maintain a log listing the exact times at which the device was removed and reapplied. The MOX3 uses two ECG leads and samples at 400 Hz. Raw ECG recordings were acquired in BDF format, converted to EDF using EDFbrowser, and then processed in Kubios HRV Scientific (version 4.1.2). During BDF-to-EDF conversion, a high-pass filter was applied in EDFbrowser to reduce low-frequency baseline drift; a 0.5-Hz cutoff was used by default. In Kubios, recordings were visually inspected, RR intervals were extracted, automatic beat correction was applied, and mean heart rate together with the time-domain HRV metrics RMSSD and SDNN were derived. Time-varying HRV analysis used 300-s windows with a 60-s grid interval. Minute-level ECG epochs were excluded when signal quality was <80%, beat-correction burden exceeded 5%, mean heart rate was <30 or >220 beats/min, HRV values were negative or >500 ms, or mean heart rate was missing. Chamber-level summaries required ≥70% clean chamber epochs and hourly summaries required >30 valid minutes per hour. For free-living wake/sleep HRV summaries, participants were included only when they contributed at least three post-chamber days with ≥360 valid minutes per day and ≥48 total clean recording hours.

#### Continuous Glucose Monitoring

Interstitial glucose concentrations were assessed using a blinded professional continuous glucose monitoring (CGM) system (FreeStyle Libre Pro; Abbott). The sensor was applied to the back of the upper arm on Day 1 and was removed upon exit from the room calorimeter on Day 3. Timestamped blood glucose time series data were downloaded retrospectively using the LibreView portal. For each participant, these timeseries were cleaned by excluding pre-chamber values and readings flagged during quality control as representing implausible spikes or likely compression-related artifacts; valid readings were then used to derive hourly glucose trajectories and chamber-period averages.

#### Skin Temperature Monitoring

While participants were in the room calorimeter, their skin temperature was measured using wearable Thermochron iButton loggers (DS1921H-F5; Maxim Integrated/Analog Devices). Five sensors were attached to the skin with Tegaderm immediately before chamber entry on Day 2 and removed immediately upon chamber exit on Day 3. Sensors were placed at the abdomen, left wrist, right calf, and left and right infra-clavicular areas. Temperature was recorded at 1min intervals using OneWireViewer software. For analysis, proximal skin temperature was derived from the infra-clavicular sensors, distal skin temperature from the wrist and calf sensors, and the distal–proximal gradient was calculated as T_dist − T_prox. Distal–proximal gradients are commonly used as an index of peripheral heat dissipation in sleep and circadian physiology ^96^. Skin-temperature summaries were calculated from valid readings only after exclusion of missing or out-of-range values; hourly chamber summaries required >30 valid minutes per hour and subject-level chamber summaries required ≥70% valid proximal and distal readings.

#### Questionnaire Package and Mood Assessments

Participants completed a REDCap-based questionnaire package on the study iPad at approximately 9:30am on day 2 shortly after entering the room calorimeter. Questionnaire-derived variables included self-reported race/ethnicity and socioeconomic items used to characterize the cohort, perceived stress assessed with the 10-item Perceived Stress Scale, physical and mental fatigability assessed with the Pittsburgh Fatigability Scale, and subjective sleep restfulness from the Pittsburgh Sleep Quality Index. Questionnaire scores were calculated according to standard instrument scoring procedures and exported as subject-level summary variables for analysis.

Momentary mood was assessed hourly during the 23-h room calorimeter stay (and more frequently between 7:00-8:00am on day 3 during the awakening response) using repeated REDCap ecological momentary assessments administered on the study iPad. At each assessment, participants rated how they felt “right now, at this moment” on 8 items: stressed, anxious, fatigued, frustrated, happy, energetic, in control, and calm. Response options ranged from “Not at all” to “Extremely” on a 0-4 scale. Negative mood and positive mood composite scores were calculated as the mean of the 4 negative and 4 positive items, respectively. These data were used to generate chamber-day mood trajectories and chamber-average fatigue and energetic ratings for downstream analyses.

#### Parent MiSBIE Cohort Data Used in Extended Data figure 2

The top-row data in **Extended Data figure 2** were drawn from the larger parent Mitochondrial Stress, Brain Imaging, and Epigenetics (MiSBIE) study ^97^ rather than from the MDEE calorimetry cohort. MiSBIE was a 2-day on-site deep-phenotyping protocol with additional home-based circadian sampling in 25 individuals with the m.3243A>G mutation and 15 individuals with a single, large-scale mtDNA mutation (see ^97^ for details). For the present manuscript, parent-cohort analyses were limited to the MiSBIE measures shown in **Extended Data figure 2**: am/fasted and pm/fed resting energy expenditure, urinary norepinephrine and epinephrine, salivary cortisol, resting heart rate, and resting lactate. Parent-cohort comparisons were performed using available-case MiSBIE data, with participants carrying pathogenic mtDNA defects analyzed as a combined mitochondrial disease (MitoD) group.

#### Resting energy expenditure and body composition in the parent MiSBIE cohort

In MiSBIE, REE was measured in the sitting position on two occasions: once in the afternoon under fed conditions, and once in the morning under fasted conditions. Measurements were obtained using a VO_2_-only device, the ReeVue Metabolic Rate Analysis System (Korr Medical Technologies), with a disposable breathing tube/mouthpiece and nose clip. For these measurements, participants remained seated in a comfortable chair, were asked to relax and breathe normally, and to remain as still as possible for 10 min. Study staff documented visible air leakage, removal of the mouthpiece, and participant movement during the test. The device estimates oxygen consumption and converted it to resting energy expenditure (kcal/day) using the manufacturer’s standard algorithm. Body composition was estimated using a 4-point bioelectrical impedance device (InnerScan PRO multi-frequency segmental wireless body composition monitor; Tanita) to derive percent fat mass and fat-free mass (FFM). We calculated FFM-adjusted REE residuals using the same log-linear regression approach as described below for the AM/fasted and PM/fed conditions.

#### Urinary biogenic amines in the parent MiSBIE cohort

Urine for catecholamine analysis was voided by participants into a disposable cup or urine hat, transferred into a container with 30 mL acetic acid preservative. In the laboratory, urine was transferred to 50-mL conical tubes, centrifuged at 1000 × g for 10 min to pellet cells, and the cell-free supernatant was aliquoted and stored at -80°C until analysis. Urinary norepinephrine and epinephrine were measured as part of a broader biogenic amine panel by UPLC-MS/MS at the Biomarkers Core Laboratory at Columbia University’s Irving Institute for Clinical and Translational Research. Samples were spiked with deuterated internal standards, proteins were precipitated, analytes were derivatized with dansyl chloride, and the supernatant underwent liquid-liquid extraction with ethyl acetate. Extracted metabolites were resuspended in acetonitrile and separated on a Waters ACQUITY UPLC HSS C18 column (2.1 × 100 mm, 1.8 μm) maintained at 40°C with gradient elution using water and acetonitrile containing 0.1% formic acid at 300 μL/min. Detection was performed in positive electrospray ionization mode with multiple-reaction monitoring on a Waters Xevo TQS mass spectrometer integrated with the ACQUITY UPLC system. For **Extended Data figure 2B**, urinary norepinephrine and epinephrine were expressed relative to urine creatinine.

#### Resting salivary cortisol in the parent MiSBIE cohort

Saliva was collected in MiSBIE using Salivette devices (Sarstedt). For each collection, participants held the cotton roll on the mid-tongue for 2–5 min without biting or excessive oral movement. Saliva samples were placed on ice immediately after collection, then were centrifuged at 1000 × g for 5 min, aliquoted, and stored frozen until analysis. Salivary steroid hormones were quantified by LC-MS/MS using an API 5000 QTrap mass spectrometer (AB Sciex). After thawing, 100 μL saliva was combined with 50 μL internal standard and 100 μL methanol/water containing zinc sulfate, vortexed, centrifuged, and the supernatant was injected into the LC-MS/MS system for analysis.

#### ECG-derived resting heart rate in the parent MiSBIE cohort

Cardiac activity in MiSBIE was recorded continuously using 3-lead electrocardiography. Three BlueSensor VL electrodes (Ambu) were placed at the right and left midline subclavicular regions and the lower left abdominal quadrant and connected to a BioNex 8-slot acquisition system. ECG was sampled at 500 Hz. During the laboratory psychophysiology session, participants were instrumented and then rested quietly before the stress protocol. ECG recordings were processed using R-wave detection, and trained research staff visually corrected R-wave marking errors and ectopic beats by interpolation when needed. Heart rate was calculated as 60 divided by the RR interval. For **Extended Data figure 2B**, resting heart rate was derived from pre-stress test baseline ECG and expressed at the subject level.

#### Resting lactate in the parent MiSBIE cohort

Resting plasma lactate was quantified in MiSBIE samples using a colorimetric enzymatic L-Lactate Assay Kit (Abcam, ab65331). Plasma samples were thawed on ice and 16 µL aliquots were diluted with 184 µL of Lactate Assay Buffer in 96-well dilution plates, which were stored at −80°C until assay. The assay was performed in duplicate across 8 plate pairs (16 plates total). Kit reagents were reconstituted according to manufacturer instructions: lyophilized Lactate Enzyme Mix and Lactate Substrate Mix were each homogenized with 220 µL Assay Buffer. A 1 mM standard solution was used on each plate. Background control wells contained pooled plasma from four MiSBIE participants (two with deletions and two with point mutations). Three inter-plate reference samples were used to normalize across plates and correct for batch effects. For the assay, 50 µL of diluted plasma was used following the manufacturer’s instructions. Concentrations were determined in duplicates using the standard regression method.

#### Quantification and statistical analyses

All analyses were performed in R within a reproducible pipeline. Except for repeated daily measures, the participant was the unit of analysis. Continuous participant-level outcomes were generally compared between groups using two-sided Welch’s t-tests. Repeated daily free-living outcomes were analyzed using random-intercept linear mixed-effects models fit by restricted maximum likelihood, with group as a fixed effect and participant as a random intercept. Associations between continuous variables were assessed using Spearman rank correlations.

To adjust energy-expenditure outcomes for body size, FFM-adjusted residuals were derived from control-based allometric models fit on log-transformed values [log(outcome) ∼ log(FFM)]. Predicted values were back-transformed to the original scale, and residuals were calculated as observed minus predicted kcal/d. This framework was applied to chamber TEE, REE, DLW-derived TEE, chamber lights-on and lights-off TEE, and sleep EE where indicated. When variables were displayed as fold-change relative to the control mean, statistical testing was performed on the underlying raw values.

For integrated chamber-day analyses, within-subject associations between hourly residual TEE and concurrent physiological variables were quantified using Spearman correlations across paired hourly values, requiring at least 10 valid paired hours per participant. Group differences in these subject-level correlation coefficients were tested using two-sided Wilcoxon rank-sum tests. For the prespecified SomaScan screening analysis only, chamber-day mean marker values were log10-transformed and compared between groups using one-sided Welch’s t-tests (alternative hypothesis: MitoD > Control), followed by Benjamini-Hochberg correction across the 21 screened markers. Missing data were handled by available-case analysis without inferential-stage imputation. Standardized effect sizes for between-group contrasts were reported as Hedges’ g where indicated.

#### Data availability

De-identified, analysis-ready input data required to reproduce **Figures 2–7** and **Extended Data figures 1–6** have been deposited under restricted access: https://doi.org/10.5281/zenodo.20330836. Because the dataset contains participant-level human data from a small rare-disease cohort, access to participant-level files is controlled and available to qualified researchers for non-identifying scholarly use by request.

#### Code availability

All original code used to reproduce **Figures 2-7** and **Extended Data figures 1-6** has been deposited at GitHub (https://github.com/mitopsychobio/2026_MDEE_paper_code) and archived at Zenodo: https://doi.org/10.5281/zenodo.20334016. The archived release is the version corresponding to this version of the manuscript.

